# CSF and PET biomarkers for noradrenergic dysfunction in neurodegenerative diseases: a systematic review and meta-analysis

**DOI:** 10.1101/2021.09.30.21264271

**Authors:** E. Lancini, L. Haag, F. Bartl, M. Rühling, NJ. Ashton, H. Zetterberg, E. Düzel, D. Hämmerer, MJ. Betts

**Author notes:** **Corresponding author:** Elisa Lancini, Institute of Cognitive Neurology and Dementia Research, Leipziger Str. 44, 39120 Magdeburg. These authors contributed equally (D. Hämmerer, MJ. Betts).

## Abstract

The noradrenergic system shows pathological modifications in aging and neurodegenerative diseases and is thought to be affected in the early stages of both Alzheimer and Parkinson’
ss diseases. We conducted a meta-analysis of noradrenergic differences in Alzheimer’s disease type dementia (ADD) and Parkinson’s disease (PD) using CSF and PET biomarkers. CSF noradrenaline (NA) and 3-methoxy-4-hydroxyphenylglycol (MHPG) as well as NA transporter availability (PET MeNER) levels in controls, ADD and PD patients was summarized from 26 articles (1025 patients and 839 controls in total) using a random-effects model meta-analysis. Compared with controls, PD patients showed significant reductions in CSF NA and MHPG, and PET MeNER binding in the hypothalamus. In ADD, MHPG levels were increased compared with controls. Age correlated with CSF MHPG levels in ADD, but not in PD. Noradrenergic dysfunction in neurodegenerative diseases can be detected using CSF or PET measures and may be more pronounced in PD compared to ADD.

## 1. Introduction

Pathological alterations to the locus coeruleus (LC), a major source of noradrenaline in the brain, occur early in Alzheimer’s disease (AD) and Parkinson’s disease (PD) (1–3).

In AD, tau accumulation in the LC precedes volume loss, with a decrease of 30-55% of LC neurons from prodromal to severe dementia (2,4). The loss occurs predominantly in the rostro/middle portion of the LC (1,2,5), and correlates with decreased cognitive functions, postmortem neuropathology (4) and reduced NA levels in the cortex and hippocampus (6). Hyperactivation of remaining neurons might further accelerate tau propagation in the brain through LC projections (7,8) leading to β-amyloid deposits (9,10). In agreement with the hypothesized hyperactivation in early stages, NA increase might contribute to anxiety and depression (11), symptoms considered risk factors for developing AD (12).

In PD and synucleinopathies, a-synuclein containing Lewy bodies and neuronal cell loss in the LC (1,2) may affect NA synthesis (13) and precede degeneration to the substantia nigra (3,14,15). In support of this, a number of different clinical features related to noradrenergic dysfunction have been observed in PD (16) particularly non-motor symptoms (17) that may precede motor symptomatology and become more prevalent with disease progression (18). Moreover, anxiety and depression, symptoms that are hypothesized to be related to LC activation and NA release (11) and risk factors for AD (12), are also common non-motor symptoms in PD (18). A further shared characteristic is the change in morphology of LC cells following chronic exposure to tau and a-synuclein, in AD and PD respectively (8). The consequent loss of noradrenergic axons, terminals, NA and dopamine β-hydroxylase (DBH) activity in LC-efferent regions is supported by studies in animals and cell cultures (8).

Despite the uncertainty of healthy age-related cell loss in LC, there is clear post-mortem evidence of an age-related increase of tau in LC that may propagate along its axons, reaching first the transentorhinal regions and then in later stages, the neocortex (9,10). This process suggests a pivotal role of LC in both age-related brain degeneration (“seeding hypothesis”, Jucker & Walker, 2012) (19) and cognitive decline, because the presence of tau, more than β-amyloid plaques, in the neocortex correlates with cognitive impairment prior to death (20). The influence of the LC on other brain areas however might be exerted both directly via NA dysregulation, or indirectly via neuroinflammation (e.g., tau propagation) or additional LC-neuromodulators such as brain-derived neurotrophic factor (BDNF) and galanin (8).

Age-related cognitive functions most affected in aging (21–24) and cognitive reserve (25–28) also depend in part on the noradrenergic system, thus maintaining the neural density of the LC-NA nuclei may prevent cognitive decline in aging (28,29). Furthermore, early clinical symptoms of neurodegenerative diseases may originate from noradrenergic dysfunction, such as sleep-wake cycle dysregulation, depression, anxiety, agitation (30–34), impaired attention and memory (8), suggesting that the integrity of the LC-NA system may be critical for tracking the progression from healthy to pathological aging (35). Taking into account all of the above, noradrenergic dysfunction occurs in healthy aging and is amongst the earliest signs of AD- and PD-like neuropathology.

To assess whether noradrenergic differences are related to pathology and disease severity in neurodegenerative disease, in this meta-analysis and systematic review we aimed to investigate the extent of noradrenergic dysfunction in AD-type dementia (ADD) and PD, using data obtained from cerebrospinal fluid (CSF) and positron-emission tomography (PET) measures of noradrenergic dysfunction.

## 2. Methods

### 2.1 Search Strategy and Selection Criteria

We searched PubMed for English relevant articles. Four authors (E.L, M.R, L.H, F.B) independently extracted the relevant information. The criteria for inclusion were: (i) the presence of CSF and PET noradrenergic biomarkers measures; (ii) clear sample composition; (iii) clear descriptive statistics of sample size and biomarker measures; (iv) clear description of methodology. In case of missing data, correspondent authors were contacted and if no additional information was provided, data points were extrapolated from the article’s plots using the online software WebPlotDigitalizer Version 4.4 (36). Reviews and articles with previously published or unpublished data were excluded. The authors assessed the quality of the evidence with *robvis* online bias tool (Supplementary Figure 1) (37).

Throughout the paper we refer to the study participants using the umbrella terms ADD, that indicates both AD-type dementia patients with evidence of AD pathology and participants whose pathology was not confirmed, and PD, that includes both idiopathic PD and PD dementia patients.

A detailed flow diagram of the literature search and exclusion criteria is reported in Figure 1. A total of 26 studies reported a suitable comparison between healthy controls and ADD/PD and adequate data (mean and SD) for the calculation of the meta-analysis (CSF=23; PET=3), while an additional 23 studies were included in another exploratory analysis to investigate the association of noradrenergic levels in CSF with age and CSF measures of amyloid and tau pathology.

**Figure 1.**
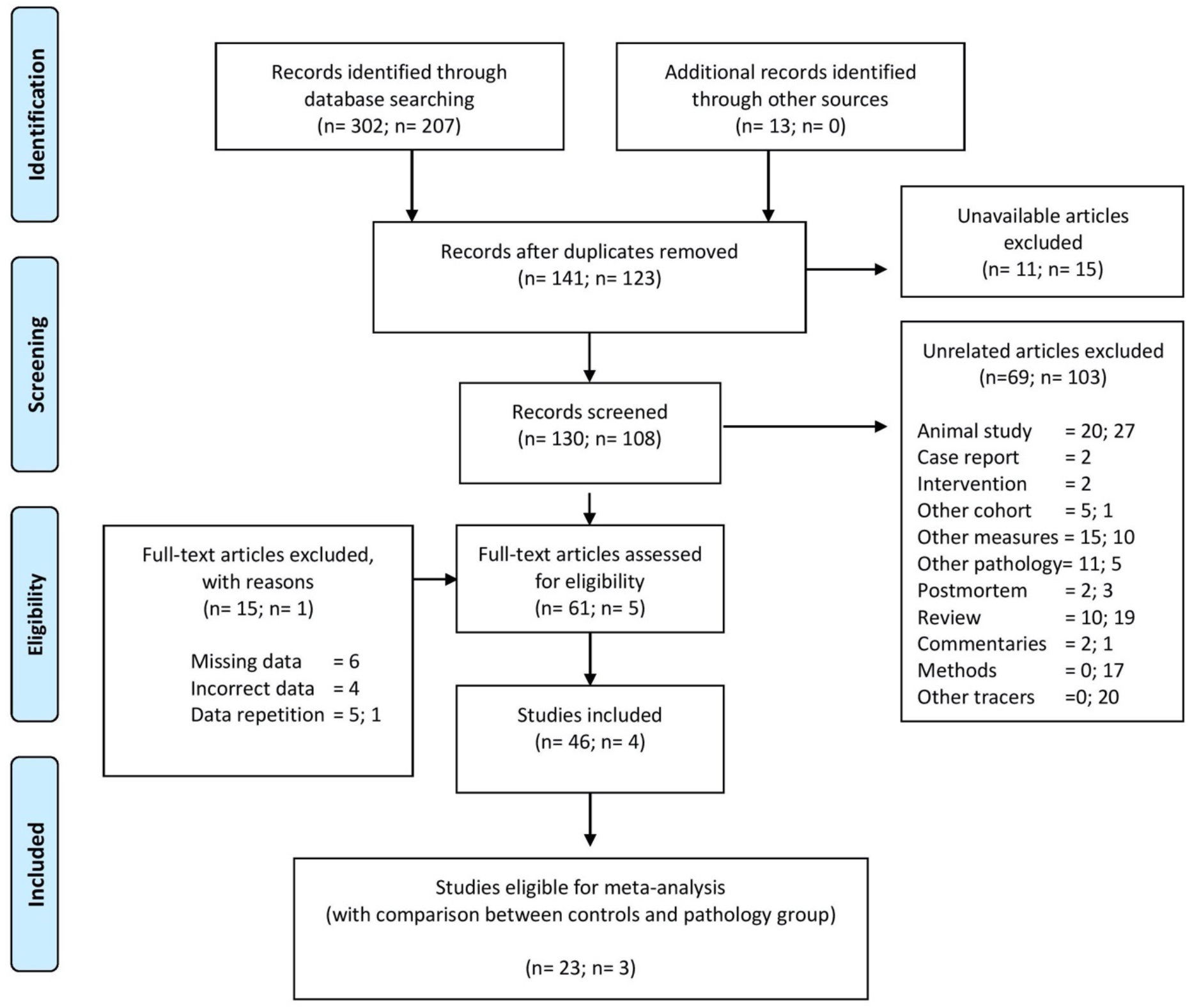
Flow diagram representing articles selection and inclusion process. PRISMA (Preferred Reporting Items for Systematic Reviews and Meta-Analyses). Figure adapted from Moher et al., 2009 (38).

Only articles included in the meta-analysis are included in the qualitative synthesis in Table 1. Studies in which the control groups were not age-matched to the ADD/PD groups were included to the extent that these studies were not outliers in the analysis and age dissimilarity was not a reported concern in the original articles.

**Table 1.**
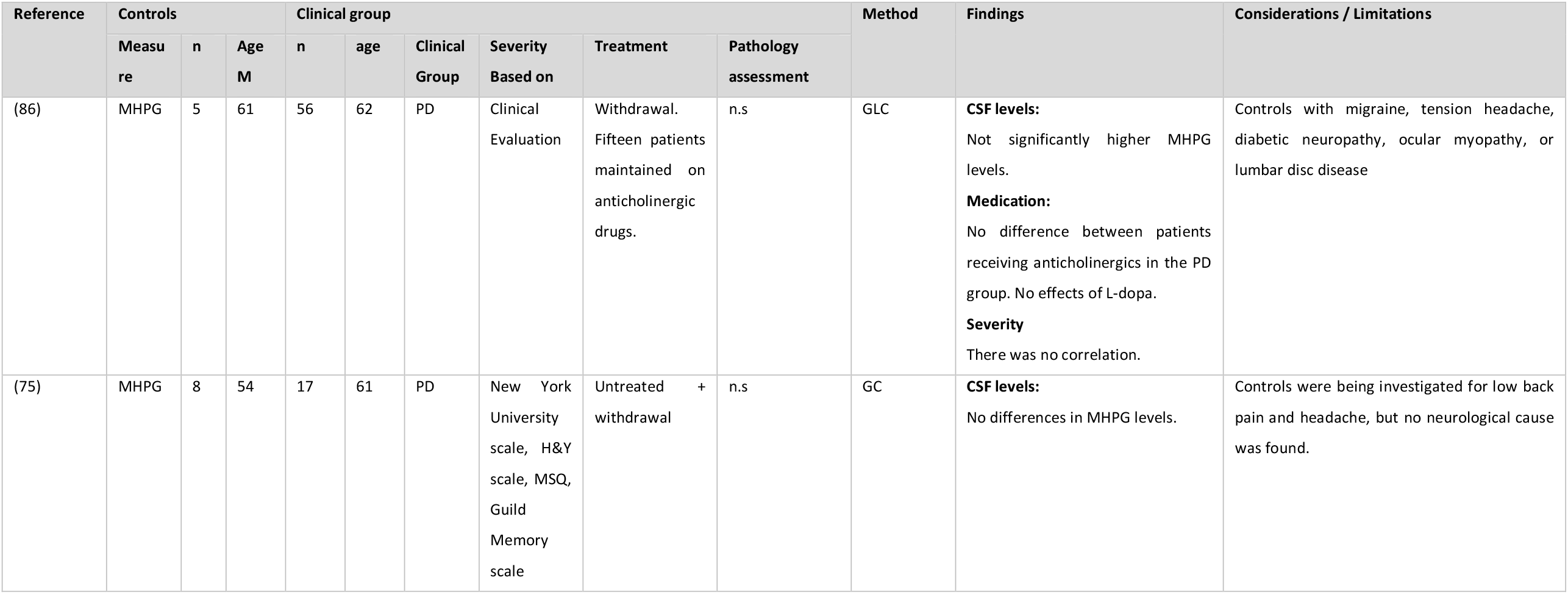
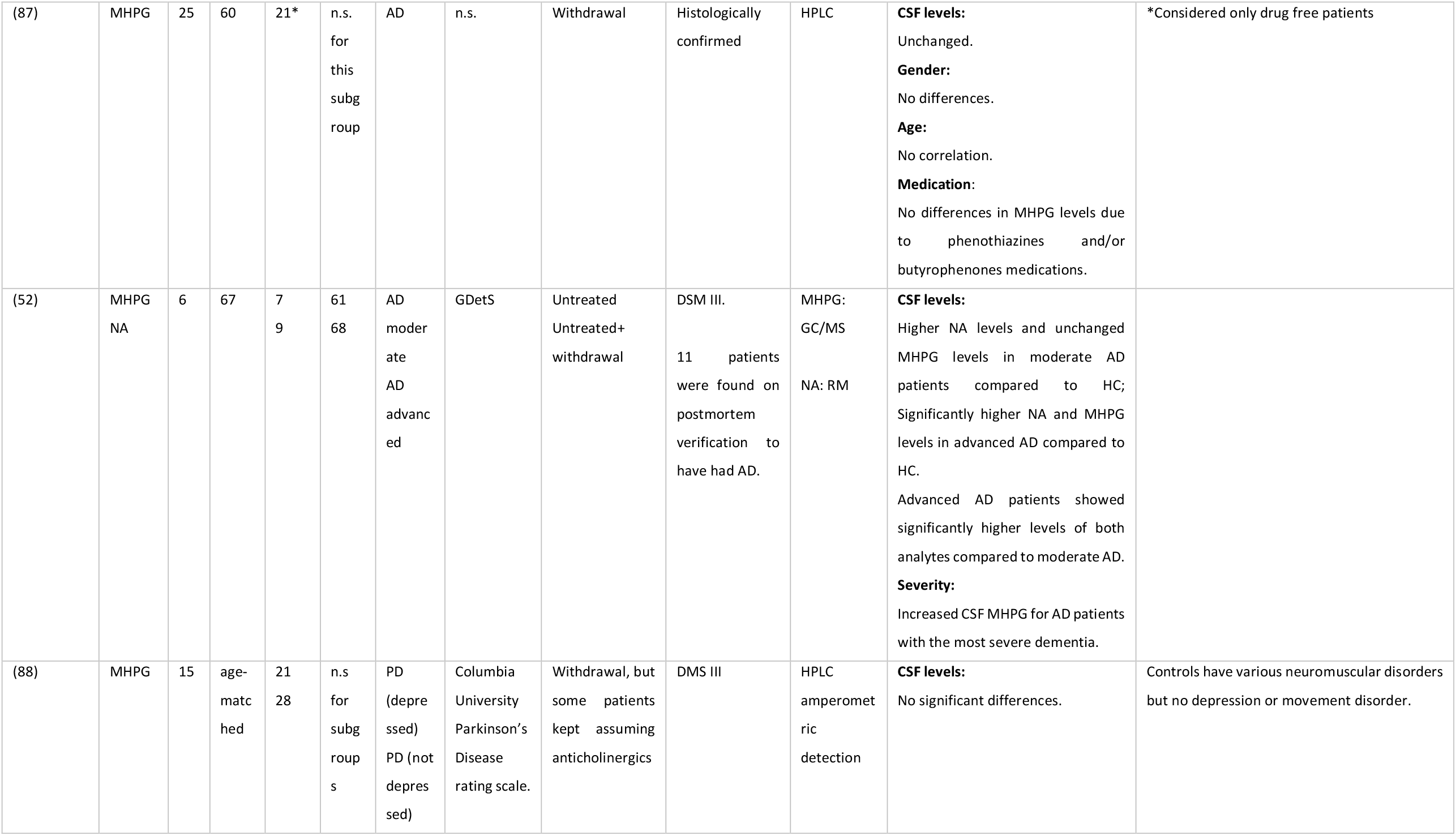
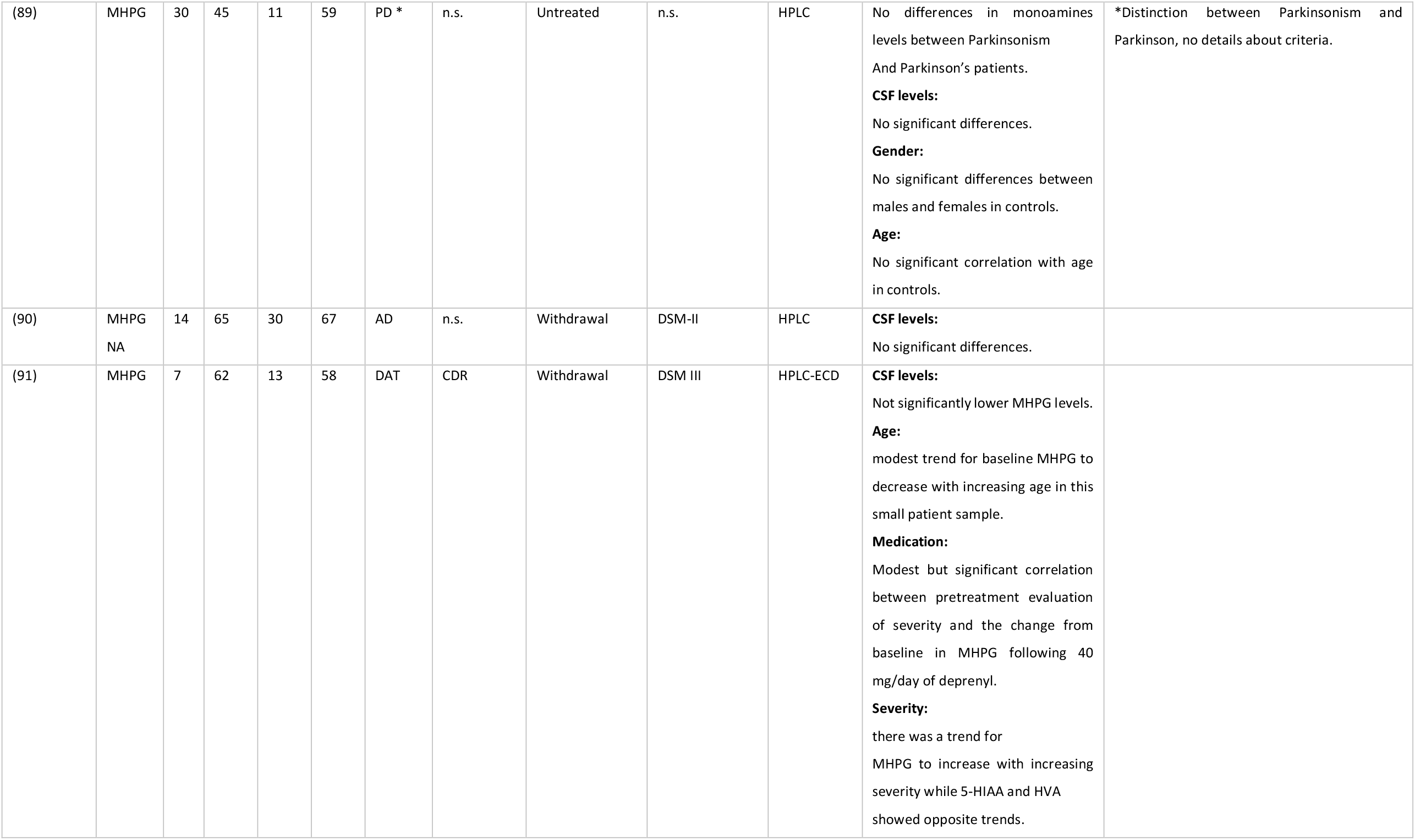
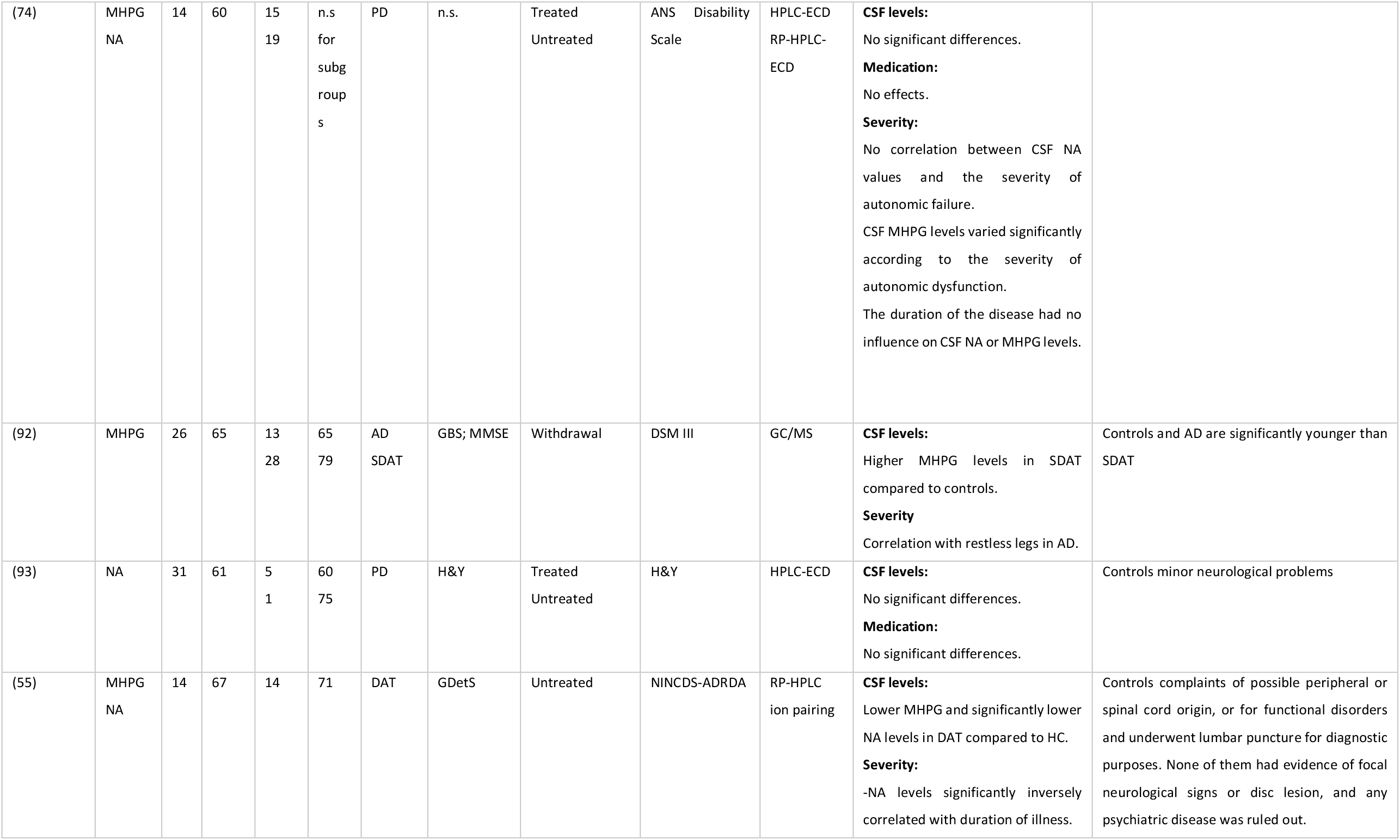
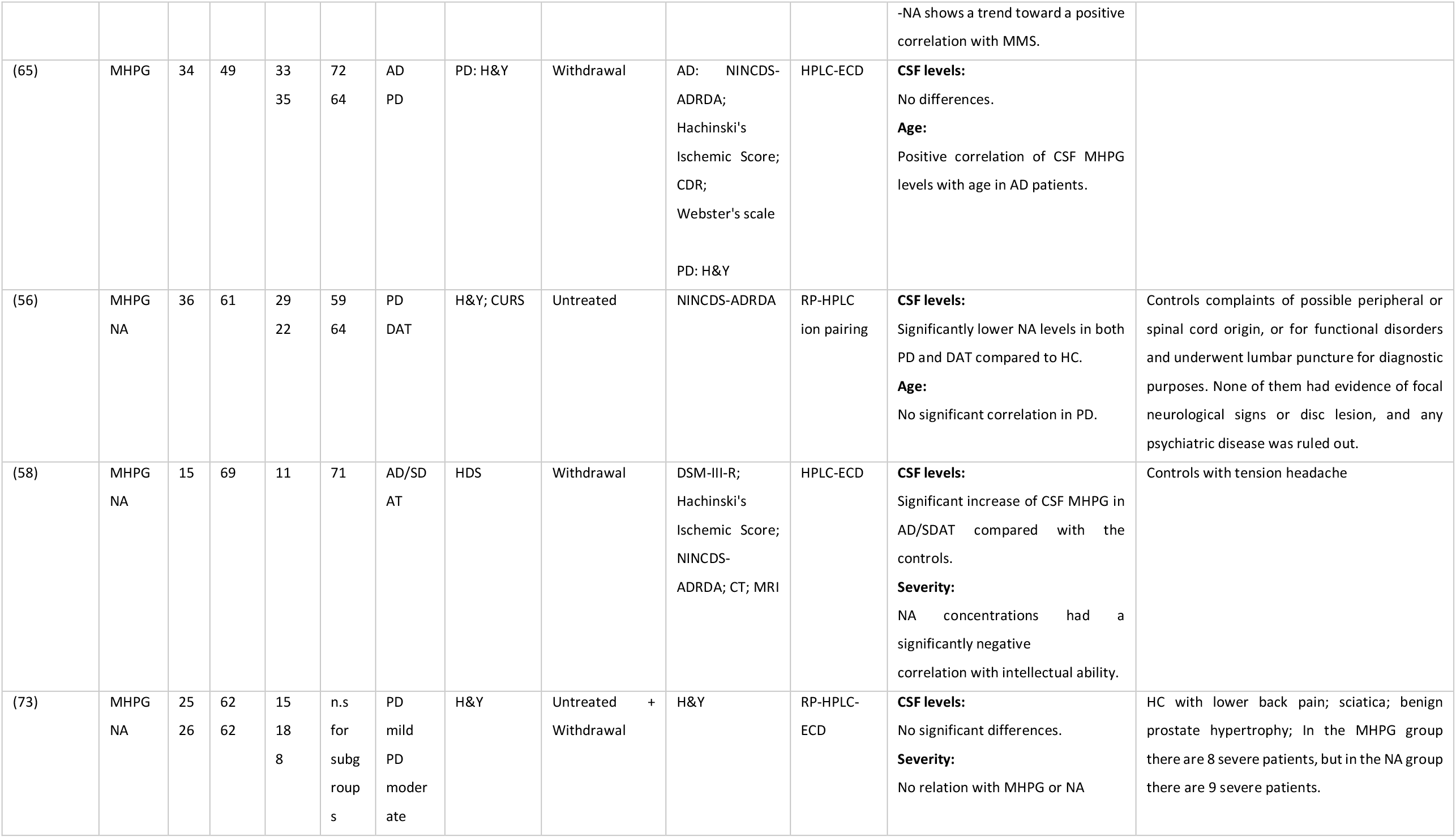
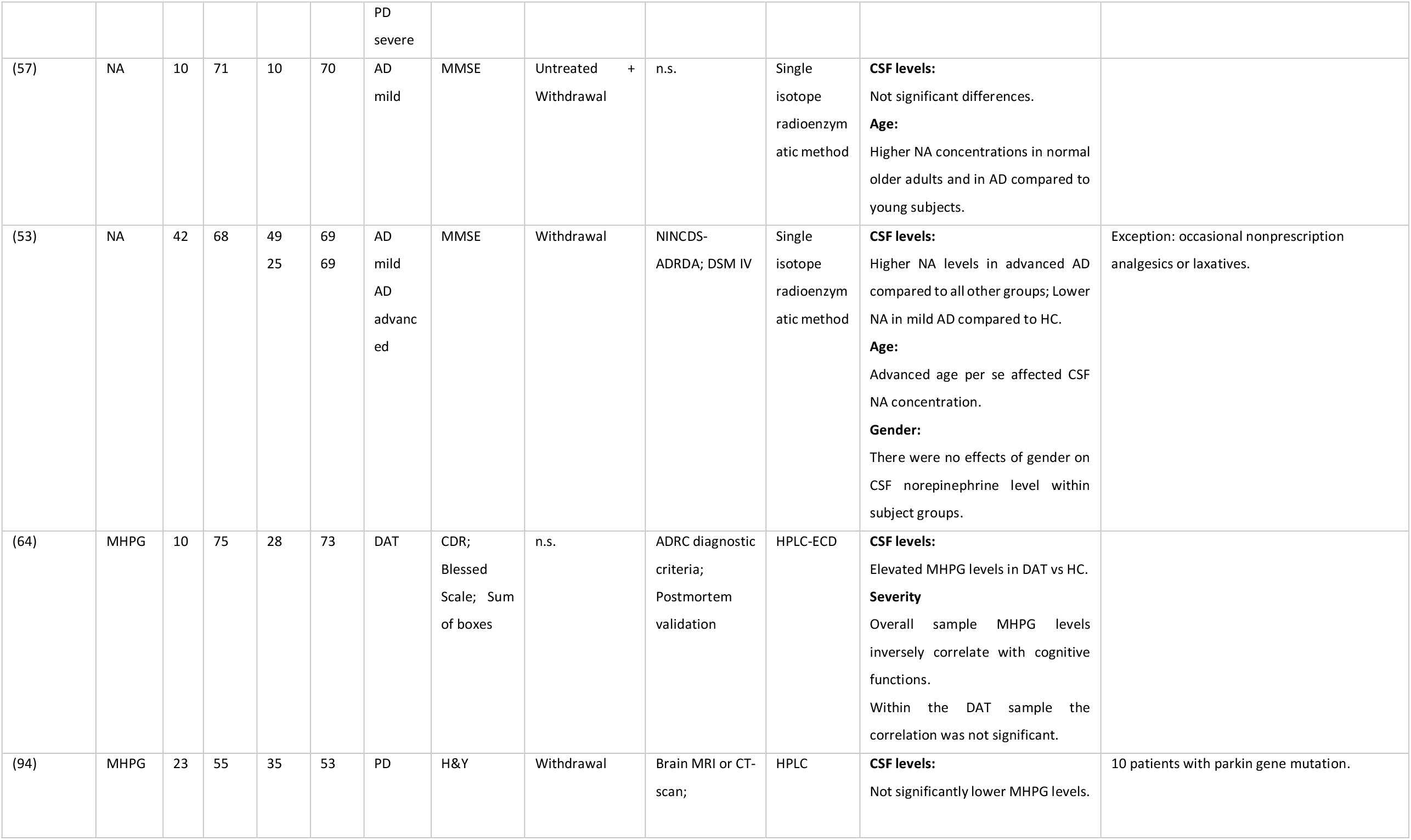
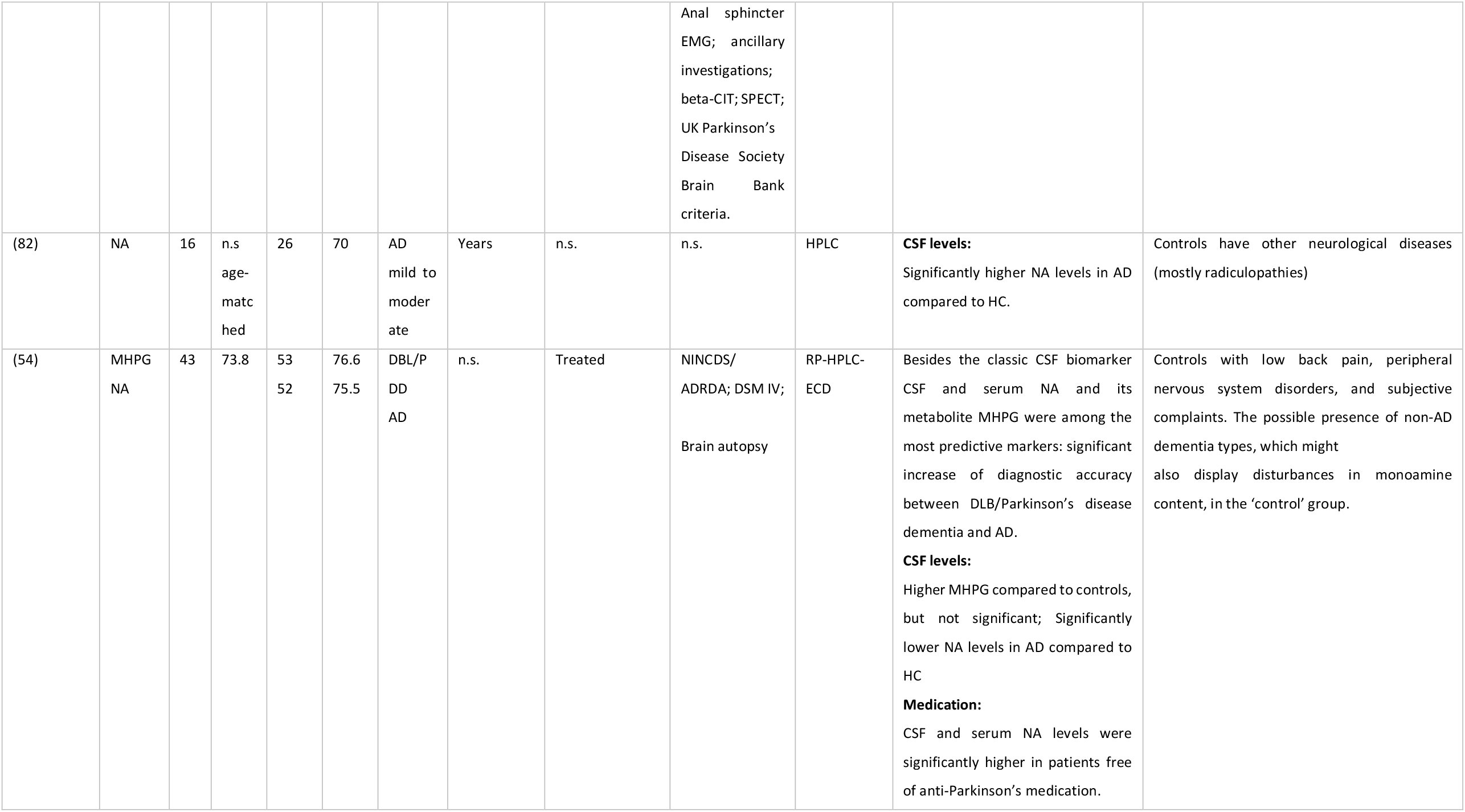
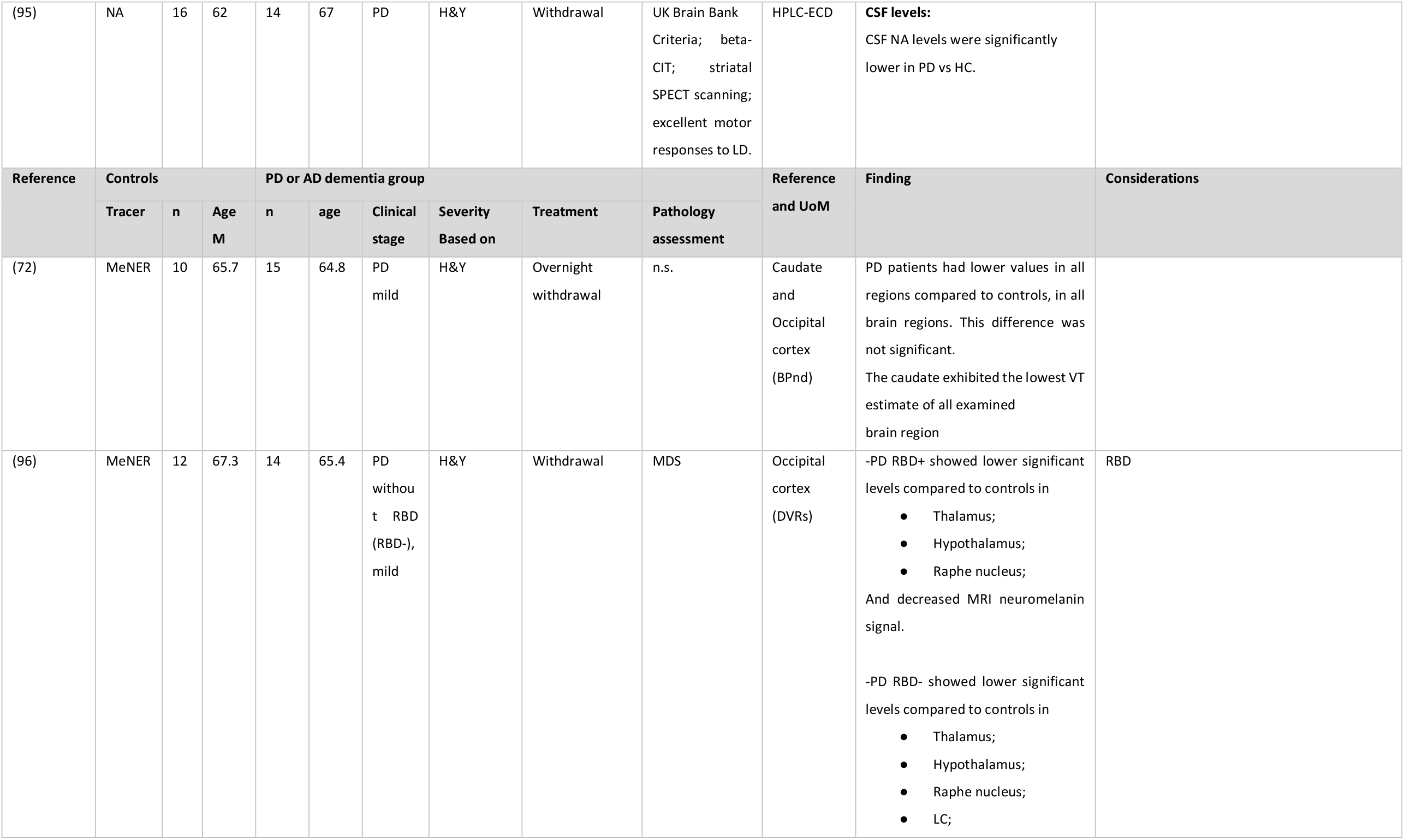
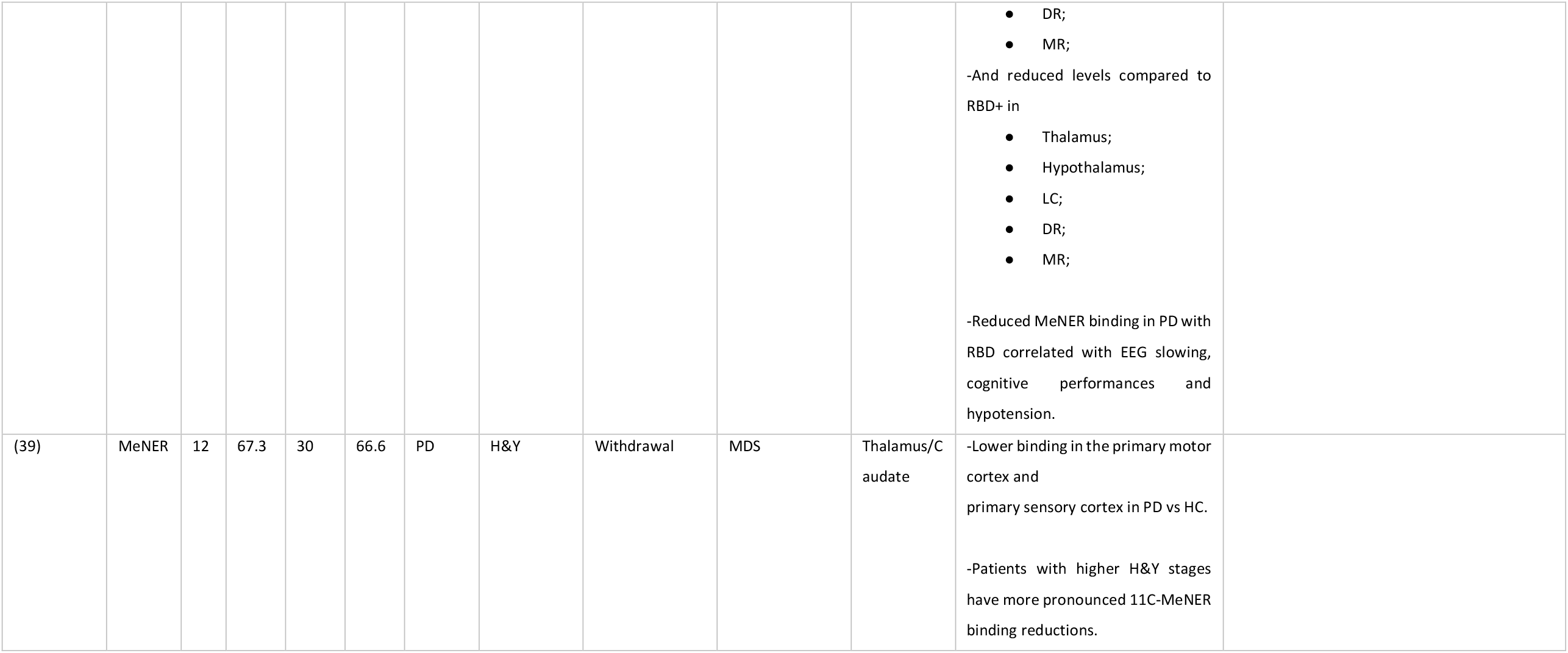
Summary of CSF (n=23) and PET (n=3) studies included in the meta-analysis. AD: Alzheimer’s dementia; CDR: clinical dementia rating; CSF: cerebrospinal fluid; CT: computed tomography; DSM: Diagnostic and Statistical Manual of Mental Disorders; ECD: electrochemical detection; EMG: electromyography; GBS: Gottfires-Brane-Stehen scale; GC: gas chromatography; GDetS: global deterioration scale; GLC: gas liquid chromatography; HC: healthy controls; HDS: Hasegawa’s dementia scale; H&Y: Hoehn and Yahr scale; HPLC: high-performance liquid chromatography; MeNER: (S,S)-11C-2-(a-(2-methoxyphenoxy)benzyl)morpholine; MDS: Movement Disorder Society; MMSE: Mini-Mental State Examination; MHPG: 3-methoxy-4-hydroxyphenylglycol; MRI: magnetic resonance imaging; MS: Mass spectrometry; MSQ: Mental Status Questionnaire; NA: noradrenaline; NINCDS-ADRDA: National institute of Neurological and Communicative Disorders and Stroke-Alzheimer’s Disease and Related disorders Association; n.s: not specified; PD: Parkinson’s disease; RBD: Rapid eye movement sleep Behavior Disorder ; RM: radioenzymatic methods; RP: reversed-phase; SPECT: single-photon emission computed tomography.

No in vivo studies using PET MeNER in ADD were found.

This review was performed according to the Preferred Reporting Items for Systematic Reviews and Meta-analyses (PRISMA) guidelines (38). Further details on the review process can be found in the Supplementary Information.

### 2.2 Data transformations

Measures of NA and MHPG in the CSF and the density of NA transporters (NETs) in the brain were extracted for all studies included in the meta-analysis. To uniform data across papers, both CSF and PET data were first transformed to mean and standard deviation measures. When data was reported using the median and interquartile range but followed symmetrical distribution, the median was used as mean and the standard deviation calculated as interquartile range *(Q3-Q1)/1*.*35*. Second, additional transformations were applied for CSF and PET data separately. CSF NA and MHPG measures were converted to the standard unit of measure, i.e., picograms or nanograms per milliliter (pg/mL, ng/mL) respectively using GraphPad. For PET studies, data were kept in the unit of measure reported by most of the papers included, namely non-displaceable binding potential (BP-ND), and when measures were reported for left and right hemisphere separately (39), the mean and standard deviation of each bilateral region was reported (40). Finally, for studies that used the same control group to compare with different ADD and PD subgroups, the number of participants in the control group was split between the subgroups to reduce the unit-of-analysis error (41). This step was not necessary for PET data because no studies reported data from both ADD and PD.

### 2.3 Random-effect mixed-model meta-analysis assessing group differences in NA levels across studies

Statistical analyses were carried out using R software (version R i386 3.4.2) (42). Only articles reporting comparisons between either ADD or PD and a control group were included in the meta-analyses. For every article, independent Welch’s t-test was conducted to assess mean differences between control and PD or ADD groups. Then, the standardized mean differences (SMD) were calculated with Hedge’s *g* correction for small samples (43). Finally, we performed a random-effect mixed-model meta-analysis using the *metagen* function from R package meta (44).

Five variables were used as explanatory variables for the meta-regression analyses: the volume of the CSF samples, the study sample size, the years post diagnosis, the laboratory analysis technique (high-performance liquid or gas chromatography, radio enzymatic methods or immuno essays) and clinical severity. Clinical severity in PD was assessed using Hoehn and Yahr (H&Y) scores in PD (mild= 1-2; moderate= 3; severe= 4-5) (45) since Unified Parkinson’s Disease Rating Scale (UPDRS) scores were not always reported. Clinical severity in ADD was assessed using Mini-Mental-State-Examination (MMSE) scores, classified according to the Alzheimer’s Association guideline (normal >24; mild= 21-24; moderate= 13-20; severe: <12) (“Medical Tests | Mental cognitive status tests | Alzheimer’s Association,” 2021).

The levels of CSF NA and CSF MHPG were investigated in both ADD (MHPG-AD; NA-AD) and PD (MHPG-PD; NA-PD), compared to controls. The levels of MeNER PET were measured only in PD compared to controls in the hypothalamus, locus coeruleus, median raphe, nucleus ruber and thalamus. The estimation of the average true effect (μ) was calculated with a 95% confidence interval and the between-study-variance using the tau-squared estimator (τ2). To control for the small number of studies included per group, we applied the adjustment method proposed by Hartung-Knapp-Sidik-Jonkman (HKSJ). The causes of the remaining statistical heterogeneity were investigated by (i) identifying statistical outliers with the R function *find*.*outliers* and (ii) potential influential cases from, studies whose exclusion from the analysis lead to significant changes in the fitted model (44). Then, we investigated studies that exert a very high influence on overall results (ii) (42) (Supplementary Figure 2). The outliers and influential cases resulting from the three analyses were then removed from subsequent analyses (Supplementary Table 1). Regarding CSF measures, a total of five studies (k=5) were identified as outliers and influential cases and subsequently removed from further analyses (NA-AD: k=2; NA-PD: k=2, MHPG-AD: k=3; MHPG-PD: k=1). The meta-analysis model was then re-calculated excluding the detected outliers and influential cases. In the PET studies, no outliers were identified and analysis of influential cases was not possible due to the small number of studies per brain region (k=2).

### 2.4 Random-effect mixed-model meta-regression

Meta-regressions were performed to assess the influence of the hypothesized explanatory variables. Using the *rma* function in *metafor* with continuous variables centered and standardized (z-scores), three regression analyses were conducted: a linear meta-regression model using clinical severity as a predictor of interest (model 1), in addition to two multiple meta-regressions models testing the interactions between the laboratory analysis technique, the volume of the CSF samples and the sample sizes (model 2), and finally taking sample size, clinical severity and years post diagnosis (model 3). The laboratory analysis technique entered the model as a between-study factor.

### 2.5 Correlation analysis of grouped data

Additional exploratory analyses were conducted on the grouped data (means and SD) to investigate the association of noradrenergic levels in CSF with (i) age and (ii) CSF measures of amyloid and tau pathology. Outliers were not excluded.

The data used to investigate the associations with age (i) did not satisfy the assumption of normality and homogeneity of variance, therefore, Spearman’s rank correlation coefficient was calculated to assess for associations between NA/MHPG levels and age, considering only articles in which the number of data points for age and CSF data were identical.

The investigation of the associations between CSF noradrenergic measures and AD pathology measures (ii) was conducted only for the ADD group and MHPG measure. In this group three articles reported both CSF MHPG levels and CSF p-tau levels whilst in the other groups MHPG CSF measures and CSF amyloid and tau measures were available only from two articles. All articles in ADD reported the same number of datapoints for CSF p-tau and CSF MHPG levels, indicating that the same number of participants were used for all measures thus Spearman’s rank correlation coefficient was calculated for all correlational analyses. There was only 1 article available reporting NA measures and CSF amyloid and tau measures, therefore no correlation analyses were conducted for these groups.

The code for all analyses described in paragraphs 2.3-2.5 are available at https://github.com/ElisaLancini/meta-analysis.

Additional analyses and the respective figures can be found in the Supplementary Material, Supplementary Figure 3, Supplementary Figure 4 and Supplementary Table 2.

## 3. Results

### 3.1 Decreased CSF and PET noradrenergic levels in PD

In PD, significant reductions in CSF NA (n= 132, g= -0.26, p=0.01) and MHPG (n= 257, g= -0.27, p=0.04) (Figure 2), in addition to reduced PET MeNER binding in the hypothalamus (n= 29, g= -0.87, p<0.05), was observed compared with control subjects (n= 114 for NA, n=184 for MHPG and n= 22 for PET respectively) (Figure 3). In the PET MeNER meta-analysis, no significant differences were observed in any other brain regions investigated (l*ocus coeruleus*: n= 22, g=-0.51, p=0.10; m*edian raphe*: n= 22, g= -0.02, p=0.95; n*ucleus rube*r: n= 22, g= -0.89, p=0.12; t*halamus*: n= 22, g= -0.95, p=0.10).

**Figure 2.**
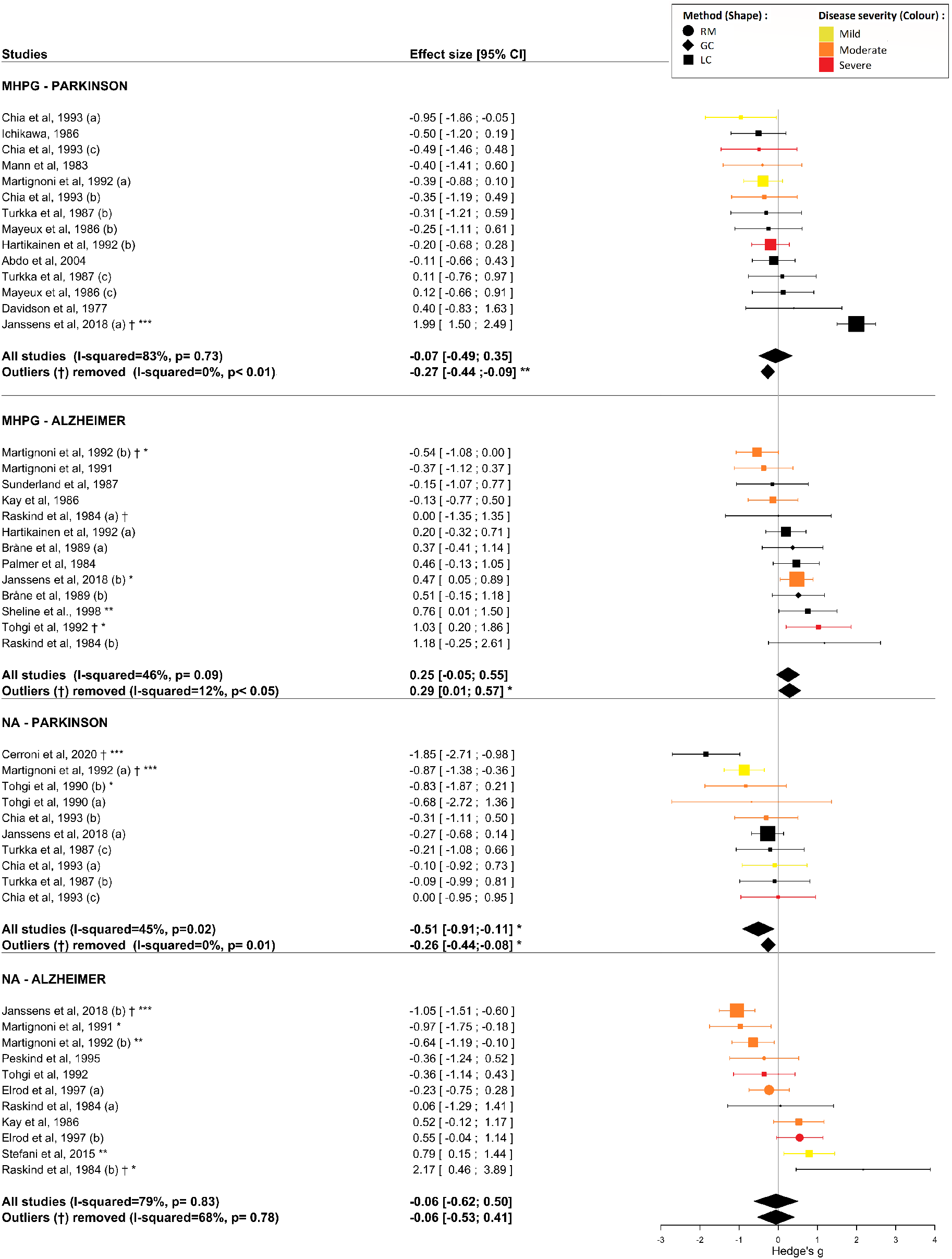
Meta-analysis results of NA and MHPG levels in CSF. The forest plot shows the effect sizes between ADD and PD compared to controls. The averaged effect size and 95% CI is indicated by the black diamonds. The size of the symbols indicates the pooled number of participants in each study. Significance levels are indicated by asterisks (*p<0.05, ** p<0.01, ***p<0.001). The significance of a single study refers to the result of the Welch’s t-test between the averages of the two groups analyzed. Studies excluded as outliers are indicated with the symbol †. The studies were characterized on the basis of the analytical method used to evaluate the noradrenergic levels of CSF, coded through the different shapes of the data points, and severity, coded through the different colors of the data points. Clinical severity was based on H&Y scores for Parkinson’s disease group (mild= 1-2; moderate= 3; severe= 4-5) and MMSE scores for Alzheimer’ dementia group (normal >24; mild= 21-24; moderate= 13-20; severe: <12). CSF: cerebrospinal fluid; GC: gas chromatography; LC: liquid chromatography; MHPG: 3-methoxy-4-hydroxyphenylglycol; NA: noradrenaline; RM: radioenzymatic methods.

**Figure 3.**
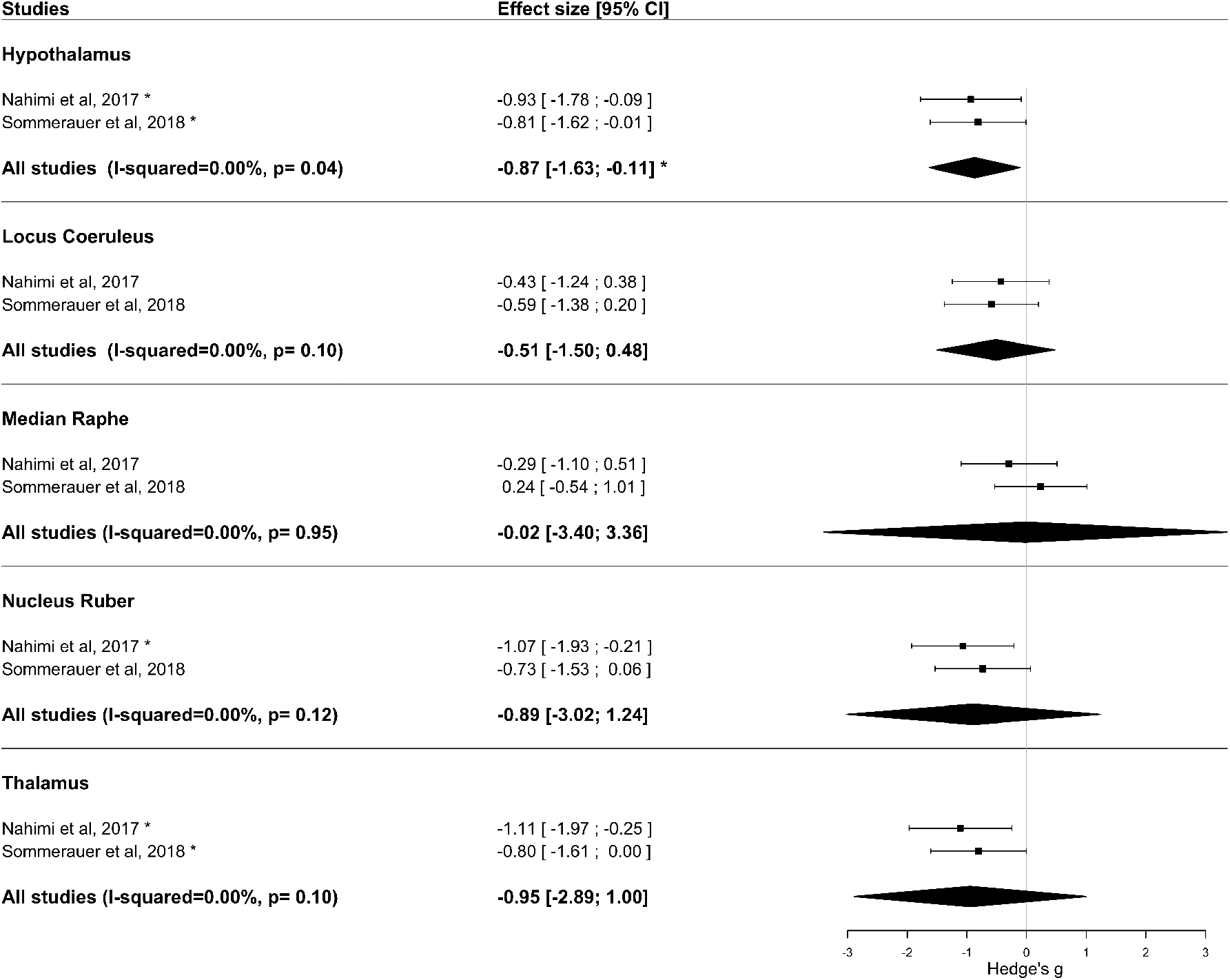
Meta-analysis results of PET MeNER binding in PD and control groups. The forest plot shows the effect sizes of the disease group compared to controls. The averaged effect size and 95% CI is indicated by the black diamonds. The application of the HKSJ method results in more conservative CI, that might exceed the variance of the single studies when the number of included studies is small and when standard errors vary considerably between them. The size of the symbols indicates the pooled number of participants in each study. Significance levels are indicated by asterisks (*p<0.05, ** p<0.01, ***p<0.001). The significance of a single study refers to the result of the Welch’s t-test between the averages of the two groups analyzed. MeNER: (S,S)-11C-2-(a-(2-methoxyphenoxy)benzyl)morpholine; PET: positron emission tomography.

### 3.2 Increased CSF MHPG levels in ADD

In ADD, no significant difference in CSF NA levels (n= 194, g= -0.06, p= 0.78) was found, however a significant increase in CSF MHPG (n= 208, g= 0.20, p= 0.04) was observed compared to controls (n= 150 for NA, n= 162 for MHPG respectively). No in vivo studies using PET MeNER in Alzheimer patients were found, therefore no meta-analysis was conducted for this group.

### 3.3 Age is a predictor of CSF MHPG levels in ADD

In ADD, age significantly predicted CSF MHPG levels (n= 337, *rs(15)*= 0.73, *p*= 0.01), however, no significant association between NA levels and age was found (n=226, rs(10)= 0.40, p= 0.19). In PD, no significant association between MHPG (n=331, rs(12)= 0.29, p= 0.31) or NA(n= 49, rs(2)= 0.80, p= 0.33) was observed. In the control group, no significant association between CSF NA (n= 49, rs(22)= 0.16, p= 0.50) or MHPG levels (n= 331, rs(19)= -0.05, p= 0.82) and age was observed.

### 3.4 P-tau181 does not correlate with CSF MHPG levels in AD

Spearman’s rank correlation coefficient showed no significant association between p-tau181 and MHPG levels (n= 130, rs(1)= -0.05, p= 1) in AD.

Only two studies assessed the associations between MHPG levels and amyloid-β 42 and t-tau in AD, and two more studies assessed the association between MHPG levels and t-tau in PD. The results of the correlations were not significant in any of the groups.

### 3.5 CSF sample volume, sample size, years post diagnosis and clinical severity do not predict noradrenergic levels in any of the investigated groups

The influence of hypothesized explanatory variables, namely the volume of CSF samples, the sample size, the number of years post-diagnosis and clinical severity, and possible interactions among them, were assessed by three meta-regression models.

In the first model (model 1), severity was included as a single predictor. This linear meta-regression model was not significant in any investigated group (MHPG-ADD n pooled= 403, R2=75.51, F(1,8)=2.75, p= 0.14), NA-ADD (n pooled= 344, R2=0.00, F(1,7)=0.44, p= 0.53), MHPG-PD (n pooled= 441, R2=0.00, F(1,11)=1.65, p= 0.22), NA-PD (n pooled=246, R2=0.00, F(1,6)=0.08, p= 0.78)).

In the second model (model 2), the possible effects and interactions between the laboratory analysis technique, the volume of the CSF samples and the sample size were assessed. The resulting multiple linear regression model was not significant in any investigated group (MHPG-ADD (n pooled= 403, R2=0.00, F(6,3)=1.33, p=0.44), NA-ADD (n pooled= 344, R2=0.00, F(6,2)=0.09, p= 0.99), MHPG-PD (n pooled= 441, R2=0.00, F(5,7)=0.81, p= 0.58), NA-PD (n pooled= 246, R2=0.00, F(3,4)=2.00, p= 0.25)).

The third model (model 3) explored the possible effects and interactions of the study sample size, clinical severity and number of years post diagnosis on noradrenergic levels in CSF (NA and MHPG). The resulting multiple linear regression model was not significant in any investigated group (MHPG-ADD (n pooled= 403, R2=0.00, F(5,4)=5.56, p=0.51), NA-ADD (n pooled= 344, R2=0.00, F(7,1)=0.37, p= 0.85), MHPG-PD (n pooled= 441, R2=0.00, F(7,5)=0.36, p= 0.89), NA-PD (n pooled= 246, R2=0.00, F(3,4)=0.07, p= 0.97)).

## 4. Discussion

Our meta-analysis and review set out to identify the extent to which CSF and PET can be used to characterise alterations to the noradrenergic system in ADD and PD independent of healthy aging.

Effect sizes of the studies included in the meta-analyses were pooled and the influence of possible explanatory variables were investigated using a meta-regression analysis. Moreover, exploratory correlation analyses were conducted on grouped data (means and standard deviations) to investigate the association between CSF NA/MHPG measures with age and CSF amyloid and tau pathology.

The results from the pooled effect sizes showed that in ADD, MHPG but not NA levels were significantly increased compared to controls, and that age is a significant predictor of MHPG levels. However, in PD, we observed a significant decrease in CSF NA, MHPG and in PET NA transport binding in the hypothalamus compared to healthy controls.

### 4.1 Hypothesized increased metabolic activity to preserve noradrenergic levels in AD

We observed no significant difference in NA and increased levels of MHPG in the CSF data of ADD participants compared to healthy controls. We will discuss them in the light of other studies that have measured NA and MHPG directly in the brain, using microdialysis or brain tissue, to provide a more comprehensive frame in which to hypothesise the relationships between these measurements and the possible mechanisms underlying these differences. In studies assessing noradrenergic levels post mortem, NA has been shown to be decreased in ADD whilst MHPG levels were higher or not significantly different compared to controls (32). However, evidence from animal studies suggest that extracellular levels may differ from levels observed in tissue (46,47) and that the efficiency of compensatory mechanisms depends on the extent of the lesion of the LC (48).

In contrast, Hughes and Stanford reported higher NA extracellular levels in LC-lesioned animals compared to sham controls (49). This evidence suggests a possible difference between tissue and extracellular NA levels (50), whereby extracellular levels (assessed using microdialysis) are maintained following the depletion of noradrenergic neurons. Thus, it is currently unclear how noradrenergic differences in CSF relate to the integrity of the LC particularly in humans.

However, the result of the study by Abercrombie and Zigmond (1989) suggests that following partial degeneration of noradrenergic terminals, the remaining terminals are still able to maintain extracellular NA concentrations at a normal level, whereas this ability might be lost at a more advanced stage of neurodegeneration (48).

Antemortem brain biopsy in AD patients showed that endogenous release of NA in the frontal cortex did not vary from controls. In the temporal cortex of the same subjects, a decrease in NA and NA uptake was observed (51) but the ration between MHPG/NA was elevated. The combination of no significant difference in NA release in the frontal cortex, and increased MHPG/NA ratio in the temporal cortex may indicate a potential biological compensatory mechanism to preserve extracellular NA levels. This could corroborate the lack of significant difference in CSF NA levels and increased MHPG levels observed in our meta-analysis for ADD patients. In the same study, MHPG levels were found not to be altered which would not be in support of this idea, however it has been suggested that the NA turnover might increase with disease progression, as Raskind and colleagues found CSF MHPG levels of severe AD patients to be significantly elevated compared to controls (52). Therefore, NA and MHPG levels likely change during disease progression. However it should be noted that other studies have reported no significant difference, decreased or increased CSF measures of NA in ADD compared to age-matched controls (53–58) and increased MHPG levels in AD have not been consistently found in the literature (REFs). Moreover, increased MHPG levels in AD have also been shown to be associated with cognitive deficits (59,60), therefore hyperactivation of the remaining neurons in response to the loss of NA neurons might elicit cognitive deterioration due to excessive NA levels which would not be in support of a compensatory mechanism (for a review see Gannon & Wang, 2019).

With respect to MHPG levels in ADD, the results across postmortem studies are heterogeneous, finding either decreased MHPG levels in hippocampus and cortical regions (61), or increased levels in caudate, hippocampus and cortex (62) and frontal regions (63), as compared to controls. In ante-mortem neocortical tissue, no significant difference in MHPG levels in ADD compared to controls has been found (51). However, in the current meta-analysis we found increased CSF MHPG levels in ADD compared to healthy controls. This result corroborates previous findings in CSF, despite the discrepancy with autopsy or ante-mortem studies. A possible explanation for this discrepancy is that NA postsynaptic receptors might become hyperactivated in an attempt to compensate for decreased NA release from terminals, thus leading to enhanced metabolism and increased MHPG levels (58,64). Moreover, our results showed that CSF MHPG levels were positively associated with age as reported in other studies (59,65). This age-related increase might reflect central increased turnover or impaired uptake, or alternatively increased metabolic activity of peripheral noradrenergic neurons, as the blood-CSF barrier does not prevent MHPG diffusion between compartments (66,67).

### 4.2 Tau and amyloid pathology measures might be necessary to identify a relationship between CSF NA/MHPG and cognitive symptoms

No associations between noradrenergic levels in ADD with clinical severity or the other hypothesized explanatory variables were found. The relation between CSF noradrenergic measures in ADD and severity remains unclear: in the study from Raskind, increased MHPG levels were found to be significant only in advanced stages of ADD (52) while in another study the increase did not correlate with the severity of ADD (58). Oishi and colleagues reported a negative correlation between CSF MHPG and MMSE across ADD and controls together, while Sheline and colleagues reported a negative correlation between MHPG and cognitive function, indicated by Sum of boxes and Blessed Information-Memory-Concentration (IMC), that did not achieve significance when the control group was removed from the analysis (53,64). CSF NA levels were reported to correlate with the severity of ADD measured with the MMSE (53,60). Reduced cognitive capacity in the presence of higher levels of NA and MHPG is in disagreement with the theories on increased noradrenergic levels as a compensatory mechanism of surviving neurons. However, it is important to consider that the relationship between noradrenergic indicators in the CSF and cognitive symptoms may not be clearly found in the absence of pathological markers.

A recent study from Jacobs and colleagues in participants with subjective cognitive decline, mild cognitive impairment and AD found that the presence of both p-tau and amyloid mediates the relationship between CSF MHPG and memory scores (59).

Another hypothesis suggests that in prodromal stages of AD, the presence of tau “pretangles” in the LC may lead to neuronal hyperactivity that is associated with the first signs of cognitive impairments. With increasing tau accumulation, cognitive decline worsens and hyperactivation promotes further propagation of tau, exacerbating cognitive decline (8). It can be hypothesized that the detrimental effects of NA might also be observed in normal aging in individuals with high tau burden since alterations in levels of NA are likely detrimental to cognition (50). Studies using CSF measures of NA/MHPG combined with measures of tau pathology might help to elucidate this relationship: an elevation of MHPG levels in the early stages might occur as result of tau accumulation induced hyperactivation of noradrenergic neurons, associated with a decrease in cognitive abilities and then at more advances stages followed by tau-related degeneration. Further investigation of noradrenergic indices during disease progression might also be useful for intervention studies assessing the potential of noradrenergic treatments, as increasing NA with uptake blockade or receptor agonist drugs in early stages might not lead to the desired therapeutic effects.

Moreover, in AD mice models, it has been shown that noradrenergic receptors are dysregulated in AD, and that their activation also elicits deteriorating effects on cognition (50). Therefore, when discussing CSF noradrenergic levels and cognitive decline, it is necessary to consider the interplay between hyperactivation, receptor dysregulation and influence of amyloid and tau pathologies.

### 4.3 Significant differences between PD and controls are observed in both CSF and PET measures

In PD, the findings of a general decrease in noradrenergic measures are consistent with the previous literature. As with AD, post-mortem studies report pathology in the LC at one of the earliest stages of PD. Neuropathological findings also indicate that substantia nigra pars compacta (SNc) degeneration is accompanied by Lewy body burden and associated loss of noradrenergic neurons in the LC (68) and that LC neuronal loss may occur earlier and in greater magnitude than that of the SNc (69). However, as in AD, the patterns of disease progression in PD are heterogeneous (70,71), and often deviate from the classical staging proposed by Braak (14).

We found overall reduced PET MeNER binding in the hypothalamus in PD compared with controls. A marked reduction in NET levels in the LC, median raphe, nucleus ruber and thalamus was also observed compared to healthy controls, however these effects were not statistically significant. These results were not expected, as we anticipated that the LC and raphe would be significantly affected due to the pathophysiology of PD. However it should be noted that the included studies assessed only the median portion of the raphe. Moreover, in the study from Sommerauer (39) a significant reduction in PD patients with rapid eye movement sleep behaviour disorder (RBD) was observed in these brain regions. However, for a better comparison with the Nahimi study, we only included the PD RBD-group, in which a MeNER binding difference was not significant. The study confirms the hypothesis that the noradrenergic system is more severely affected in patients with RBD (72).

Decreased CSF NA and MHPG levels in PD compared to their control groups are consistent with the increased prevalence of noradrenergic-related symptoms that occurs with ensuing disease progression. Noradrenergic deficits are implicated in the occurrence of the non-motor symptoms in PD (17) such as cognitive impairment, sleep disorders, autonomic dysfunction, that may precede motor symptomatology and become more prevalent with disease progression (18).

In PD, CSF NA and MHPG levels do not seem to correlate with disease severity assessed using H&Y scores (73) or clinical evaluation (74). In line with the literature, the present meta-analysis revealed that H&Y scores did not explain variation in CSF NA and MHPG effect sizes between PD and control groups. It should be noted however, that the lack of association reported could be due to the use of the H&Y questionnaire: the UPDRS and more recent MDS-UPDRS would be preferred as a more sensitive measure to assess motor and non-motor symptoms. CSF MHPG levels alone are associated in other studies with the severity of autonomic dysfunction measured with the ANTS Disability Scale (74) and to be inversely correlated with performance on the digit span test (75).

### 4.4 More studies are required to determine the relationship between brain pathology and CSF NA/MHPG biomarkers

In additional exploratory analyses, we investigated the relationship between CSF NA/MHPG levels and pathology biomarkers, namely amyloid, t-tau and p-tau in both ADD and PD. This analysis was possible in the ADD group, however only three studies reported both p-tau181 and MHPG levels. The relation between p-tau181 and MHPG across these three studies was not significant. Amyloid and t-tau were only specified in two studies, therefore the analysis was not reported. Whilst the relationship between p-tau181 and MHPG was not significant in our sample, one of these studies did find that higher CSF MHPG is positively associated with p-tau, independent of amyloid (23). In the same study, MHPG was shown to be associated with memory deficits via both tau and inflammation-mediated amyloid pathology, and with neuropsychiatric symptoms through a direct pathway partially mediated by tau (23). The close relation between the NA-system and AD-related pathology has been confirmed in an animal study showing the direct etiological role of NA in AD pathology (76). The authors recreated the AD-related increase in *α*2A-adrenergic receptors activity in two independent AD mouse models and found that nanomolar concentrations of amyloid-β oligomers bind to aplha2A-adrenergic receptors leading to hyperphosphorylation of tau and increased response sensitivity of aplha2A receptors to amyloid-beta oligomers. Thus, NA could be involved in mediating the toxic effects of amyloid-β (77). Tau pathology has also been suggested to be facilitated by alterations to the LC-NA system as demonstrated in rodents (7). Moreover, whilst no significant relationship between NA/MHPG and tau was observed in our analyses, MHPG more than NA may be related to p-tau as animal studies have shown that NA O-methylation in MHPG is necessary for tau spreading (78).

The lack of significance in the relationship between CSF NA biomarkers and brain pathology in our study may be attributed to the small number of studies and study heterogeneity available to us to conduct a meta-analysis but we must also consider that results obtained from animal studies may not translate to humans, and that causal associations between AD pathology and NA dysfunction remain to be determined. Larger longitudinal studies with CSF and PET measurements are needed to shed more light on this potential relationship.

## 5. Study Limitations

### 5.1 Duplicate data

Studies using identical databases were removed when duplicate samples were reported, however the origin of samples from 41 studies was not reported so additional duplicate data cannot be fully excluded.

### 5.2 Limited data availability

Only studies whose mean and standard deviation was provided or was calculable from other descriptive measures were included in the study, and not all studies reported data in this format required to calculate effect sizes.

Furthermore, data from the variables amyloid, tau, disease severity, number of years post-diagnosis, CSF volume, may be under- or overestimated because the number of subjects with CSF NA/MHPG data did not all report information on whether these additional covariables were investigated.

Additionally in the PD group, disease severity was typically reported using the H&Y scale, however the UPDRS or MDS-UPDRS would have been more desirable measures to define motor and non-motor symptom severity.

### 5.3 CSF sampling and analytical method

The volume of CSF for each measure was not always specified thus it was not possible to control for possible biases in the gradient of concentration. The volume of CSF taken can influence the concentration of biomarkers, for two reasons. First, small CSF samples will reflect the composition of the lumbar dural sac, while larger samples may also represent the rostral spinal or ventricular CSF. In addition, different molecules have a characteristic rostrocaudal concentration gradient (79). However, this may only be relevant for NA measures since previous studies showed that MHPG levels in lumbar CSF did not differ from ventricular CSF (52,80) and across fractions (81).

Although we tried to account for differences in collection and analytical techniques, there may be further differences within the same CSF analysis method, due to between and within-lab biases.

### 5.4 Pathology sample characterization

Among the ADD studies included in the meta-analysis, CSF amyloid and tau biomarkers, or confirmation of AD pathology post mortem was performed in only 15% of the studies included in the analyses presented while the majority used symptom-based scales or cognitive tests to assess disease severity. Moreover, the duration of the disease was calculated either as years post diagnosis or in some cases years from first visible symptoms reported by family members.

### 5.5 Control sample characterization

Among the included studies reporting NA levels in AD, control subjects in four studies were reported to have other comorbidities (55,56,58,82). Moreover, despite neurological and psychiatric problems being ruled out, other diseases for which controls were hospitalized might have influenced noradrenergic levels. The stress (83) caused by hospitalisation may have also influenced the results reported in our meta-analysis.

Moreover, despite the control group not reporting the presence of other pathologies, CSF NA levels increase with age (84) and the first pathological changes of AD emerge decades before the appearance of cognitive symptoms (85), thus the presence of preclinical AD in the control groups cannot be excluded.

### 5.6 Blood-brain and blood-CSF permeability of NA/MHPG

In contrast to NA, MHPG rapidly diffuses through the blood-brain (66), and blood-CSF barriers (67). Thus CSF MHPG levels might not directly correlate with central noradrenergic metabolism (66). In this respect, we should be prudent about indicating it as a pure index of CNS levels and interpreting results as such.

### 5.7 Medications

When data was reported separately based on the medication status of the participants, we included these participants in separate subgroups. However, sometimes this was not possible because the data were reported for the entire sample without confirming their medication status.

The majority of the studies reported no difference among medicated vs unmedicated participants.

## 6. Conclusion

In this review and meta-analysis, we provided an overview and quantification of noradrenergic differences in aging, ADD and PD, measured in CSF and PET. Our meta-analysis revealed a general decrease in noradrenergic levels in PD, measured as noradrenergic CSF levels and PET NA transporters availability in the hypothalamus. In ADD, an increase in CSF MHPG levels was observed which was found to be significantly predicted by age. Furthermore, explanatory and potential confounding variables were also assessed in the small subgroup of studies that reported them, and found not to influence NA/MHPG group comparisons across studies. In this regard, the discussion on possible biases of previous measurement procedures will be a valuable aid for the development of an optimized protocol for sampling and analysis of CSF measures of noradrenergic dysfunction.

This meta-analysis contributes to a better understanding of the noradrenergic system in aging, ADD and PD and demonstrates CSF measures of noradrenergic function may be differentially affected in ADD and PD. However, further studies using longitudinal CSF and PET measures in both pathology assessed AD and PD patients are required to elucidate the relationship between noradrenergic dysfunction and the progression of AD and PD. Accurate knowledge of the pathological differences of the noradrenergic system, detectable by CSF and PET, would be beneficial in a clinical setting. The characterisation of pathology-specific trajectories would contribute to the future development of a biomarker-based diagnostic model for neurodegenerative diseases and the possibility to monitor the status of the noradrenergic system might aid in early detection of pathological decline and be useful for determining the efficacy of NA drugs in clinical trials.

To support our conclusion, further studies should investigate pathologically and cognitively characterised, medication-free patients and healthy, pathology-free, age-matched control groups with PET and CSF measurements, using an optimised and shared protocol for CSF sampling.

## Data Availability

The data used for this meta-analysis are partly openly accessible by consulting the articles from which they were extracted, and partly sent by the authors after our specific request. The codes used to conduct the meta-analysis are available on the GitHub page of the corresponding author.

https://github.com/ElisaLancini/meta-analysis

## 9. Acknowledgements

We thank Calida Pereira for the assistance with R scripting, particularly with those needed to generate the figures, Valentin Baumann for providing useful suggestions on the meta-regression models and Eleonora Cuboni for helping with the conversion of CSF data.

