## Supplementary information for "CSF and PET biomarkers for noradrenergic dysfunction in neurodegenerative diseases: a systematic review and meta-analysis"

**CONTENTS**

Pages 2-3	Supplementary Methods

Pages 4-5	Additional Analyses

Page 6	Supplementary Figure 1. Risk bias assessment

Pages 7-9	Supplementary Figure 2. Influence analysis and Graphic Display of Heterogeneity (GOSH) plot analysis

Page 10	Supplementary Figure 3. Forest plot of MHPG levels (Means and SD) in CSF

Page 11	Supplementary Figure 4. Forest plot of NA levels (Means and SD) in CSF

Page 12	Supplementary Table 1. Meta-analysis results

Page 13	Supplementary Table 2. Weighted averages

Pages 14-15	Supplementary References

### Supplementary methods

**Keywords:**

We searched PubMed for English relevant articles using the keywords “CSF noradrenaline”,

“PET noradrenaline”, “PET noradrenergic”, “CSF noradrenergic”, “PET MeNER”' together with

“Parkinson”, “Alzheimer”, “dementia”, “ageing”, “aging”, “neurodegeneration”.

**Search strategy:**

Three authors (E.L, M.R, L.H) reviewed and selected the articles based on the abstracts. When there was disagreement between the selected studies, the authors reached a consensus via

discussion. The enrolled studies were then reviewed for inclusion criteria by E.L.

**Selection criteria:**

The literature search in PubMed ended in January 2021 and resulted in 509 articles (CSF= 302; PET= 207). An additional 13 articles were identified in the references of these articles

identified on PubMed. After duplicate removal, the remaining 264 articles (CSF= 141; PET=

123) were screened and a further 26 articles were excluded since they were unavailable

online (CSF= 11; PET= 15). An additional 172 articles (CSF= 69; PET= 103) were excluded as unrelated to the purpose of this review (i.e. animal studies, studies not in ADD or PD or studies not related to the noradrenergic system). From the remaining 66 articles (CSF=61; PET= 5), 16 were excluded because (i) data was missing, (ii) not reported as mean and standard deviation or in a format that did not allow for transformation into mean and standard deviation (iii) or because the reported data was already present in other studies included in the meta-analysis. In the event that the same sample was shared across studies, we included the study that was considered to be most relevant to the scope of the meta-analysis. Data from the remaining 50 studies (CSF=46; PET=4) was collected and any missing data was requested by contacting the authors of articles eligible for the meta-analysis. Finally, 26 studies reported a suitable comparison between healthy controls and ADD/PD and adequate data (mean and SD) for the calculation of the meta-analysis (CSF=23; PET=3), while the other 23 were included in another exploratory analysis to investigate the association of noradrenergic levels in CSF with age and CSF measures of amyloid and tau pathology.

Studies in which the control groups were not age-matched to the ADD/PD groups were

included to the extent that these studies were not outliers in the analysis and age dissimilarity

was not a reported concern in the original articles.

**Database control:**

The database control revealed that studies from Abdo 2007, Chia 1995, Raskind 1999 and Tohgi 1990 and Mayeux 1984 (1–5) used the same database as Abdo 2004, Chia 1993, Peskin 1995, Tohgi 1993 and Mayeux 1986 respectively (6–10). The former articles were excluded and the latter retained as they reported either the same number of subjects (8), a higher number of subjects (6,7), or more information(4,10) compared to the articles using the same subjects. Freed, 1989 and Mayeux, 1986 used the same control subjects (10,11), thus only Freed, 1989 (11) is reported in the overall analysis.

No ante-mortem studies using PET MeNER in ADD were found.

### Additional Analyses

The data collected for this review and meta-analysis is divided into two separate datasets.

The first (dataset 1) contains studies for which the calculation of effect sizes for between group differences was possible. The second dataset contains means and standard deviation data from studies included in the meta-analysis in addition to those in which a calculation of the effect size between groups was not possible, i.e., where data was only reported from one of the investigated groups (dataset 2).

To assess absolute differences in CSF measures across studies and between groups (HC; ADD; PD), data from this second dataset was used to create a forest plot of means and standard deviations (Supplementary Figure 3, Supplementary Figure 4). The arithmetical averaged mean was calculated for each group (HC; ADD; PD) and measure (MHPG; NA), and a value of 2 standard deviations from the arithmetical averaged mean was used to detect outlier data in every group. Studies identified as outliers were removed from subsequent analyses. An averaged mean and SD were calculated for each group (HC; ADD; PD) and measure (MHPG; NA). Differences in the averaged mean between groups were calculated using the Kruskal-Wallis test. This analysis was not conducted for PET studies as the groups (PD; HC) were composed of only two studies. For the data visualization plot, weighted averaged means of the groups were calculated using the sample size as weight (Supplementary Table 2).

The analyses did not show any significant difference, thus are not in line with the meta-analysis results that showed increased MHPG levels in ADD and a general noradrenergic decrease in PD. This discrepancy might be due to various factors. As the meta-analysis on dataset 1 was based on effect sizes in group differences, while the analysis on dataset 2 was based on absolute group-specific values, more significant results in the difference-based analysis could be due to a positive publishing bias. Also, heterogeneity of samples and methods should be more pronounced in datasets that are not based on difference scores. Heterogeneity in results between studies may be due to several reasons such as different inclusion criteria, for example the inclusion of "age-matched" control subjects was not always considered; the inclusion of controls who were considered healthy, but might suffer from other diseases (Martignoni et al., 1991, 1992: hospitalized for diagnostic purposes, Stefani et al., 2015: radiculopathies, Tohgi et al., 1992: tension headache) (12–15), in which an influence on the noradrenergic system could not be totally ruled out. For example, in the study by Tohgi and colleagues (15), tension headache was not considered an exclusion criteria for healthy participants. However a more recent study hypothesised an association between tension headache and dysfunctional noradrenergic pain modulation (16). Thus in this case, tension headache might have introduced a bias in the comparison between healthy and clinical groups. Moreover, the absence of pathological confirmation makes this point one of the most likely sources of bias for the standardization of the samples across studies. Finally, differences in sensitivity and specificity of CSF analytical techniques used between studies and subsequent development of these techniques over the years may have influenced comparisons across studies. Differences in CSF noradrenergic levels within studies may also be present and affect the comparability of results even within the study itself, as other medications besides levodopa (ADD: Palmer et al., 1984; PD: Turkka et al., 1987, AD and PD: Janssens et al., 2018) (17–19), were also discussed by Janssens et al., (2018) (19) to be a possible cause of bias in their results. Finally, our comparison of group measures summarized across studies (dataset2) might also have been methodologically more prone to be influenced by heterogeneity in sample sizes, as it is not possible to calculate weighted means accounting for sample sizes in the Kruskal-Wallis test. Of course, difference-based and absolute value-based meta-analyses should ultimately yield the same results.

**Supplementary Figure 1.** Risk bias assessment.

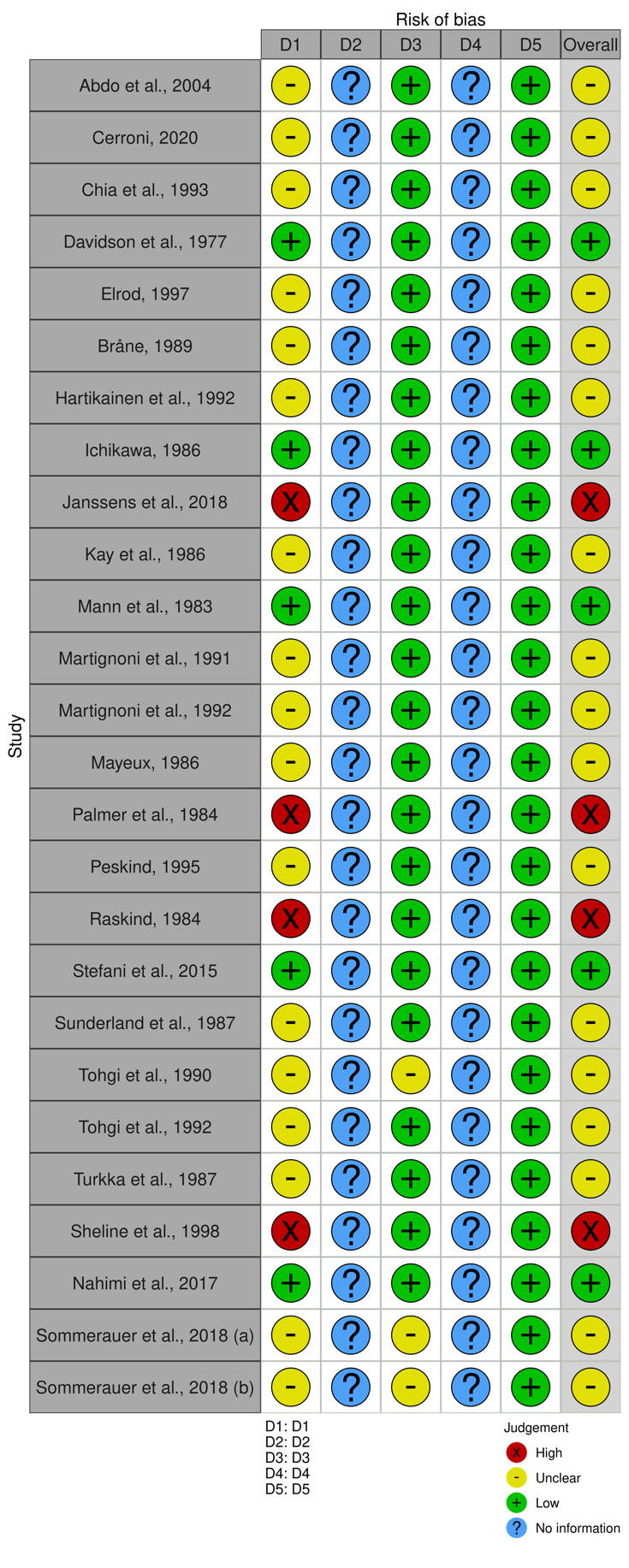

D1: bias arising from the randomization process (pathology characterization); D2: bias due to deviations from intended interventions; D3: bias due to missing outcome data (data reported in the paper or extracted from plots); D4: bias in the measurement of the outcome (information on CSF analysis method or PET tracer used); D5: bias in the selection of the reported result.

**Supplementary Figure 2.** Influence analysis and Graphic Display of Heterogeneity (GOSH) plot analysis. The figure describes the process used to detect the patterns of effect sizes and heterogeneity in the data after classical outlier removal based on standard deviations.

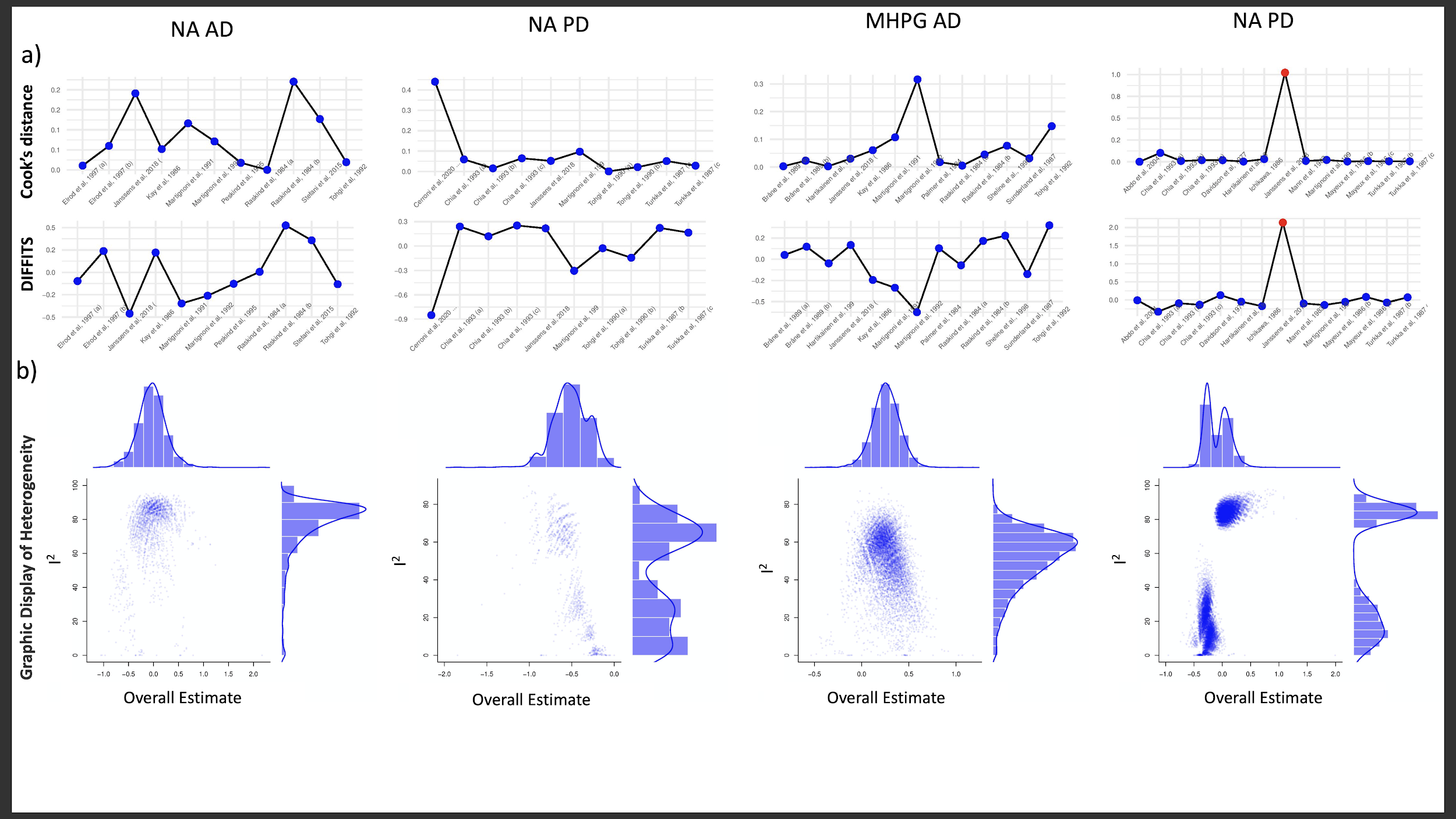

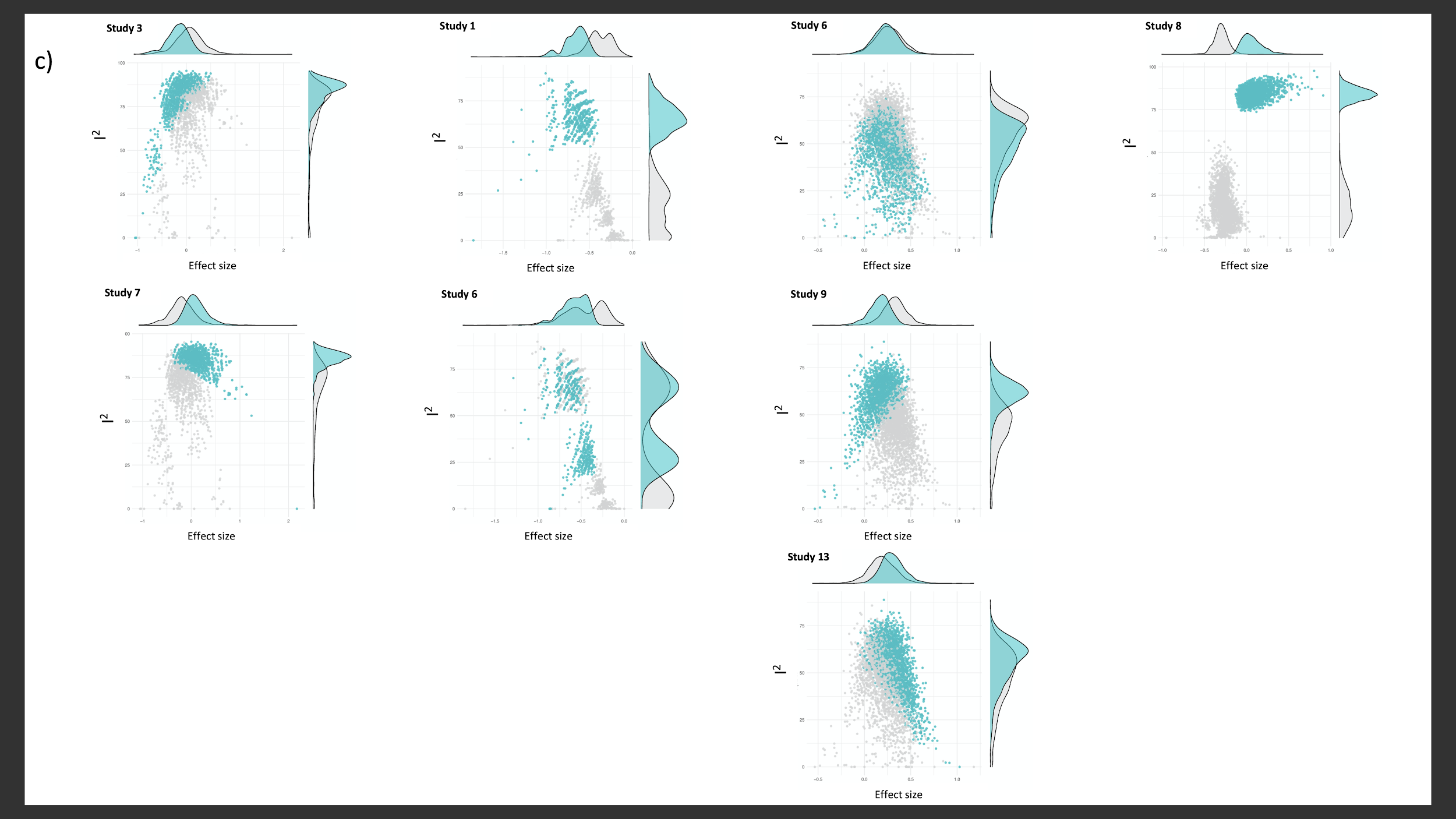

(**a**) Influence analysis with *InfluenceAnalysis* R function. The function is based on the Leave-One-Out method and recalculates the results $K-1$ times, each time leaving out one study. It assesses possible distortions in the pooled effects by detecting studies which influence the overall estimate the most. The output are plots showing the differences with and without the inclusion of each study. Here we reported the DIFFITS value and the Cook’s distance plots. These measures indicate respectively (i) how much, in terms of standard deviations, the predicted pooled effect changes after excluding this study and (ii) the distance between the values once the study is included or excluded. Values in red were identified automatically as outliers using the cut off proposed by Viechtbauer & Cheung (20). (**b**) The Graphic Display of Heterogeneity fits the meta-analysis model to all possible subsets ($2^^{k-1}$ possible study combinations). The results are a plot displaying the pooled effect size on the x-axis and the between-study heterogeneity at the y-axis, allowing us to look for specific subclusters in our data. Symmetrical distributions indicate that the effect sizes in our sample are homogeneous, asymmetrical distributions and peaks indicate the presence of subclusters (e.g in the NA PD group two peaks indicate two subclusters, that are found to be driven by study n°1 and n°6). The between-study heterogeneity (I-squared) is the percentage of variability in the effect sizes, not caused by sampling error. (**c**) Using the *gosh.diagnostics* function, three clustering algorithms (k-means, DBSCAN, Gaussian Mixture Model) detect clusters in the GOSH plot data and which studies contribute to create them. The studies reported are subsequently excluded from analyses.

**Supplementary Figure 3.** Forest plot of MHPG levels (Means and SD) in CSF. Forest plot of means and standard deviations data originated from all articles reporting data points, regardless of their inclusion or exclusion in the meta-analysis. For each study, all datapoints reporting CSF NA or MHPG levels are included. Weighted averaged means of the groups were calculated using the single studies sample sizes as weight. The averages of the weighted means are indicated by the diamonds. CSF: cerebrospinal fluid; MHPG: 3-methoxy-4-hydroxyphenylglycol; ng/mL: nanograms per milliliter.

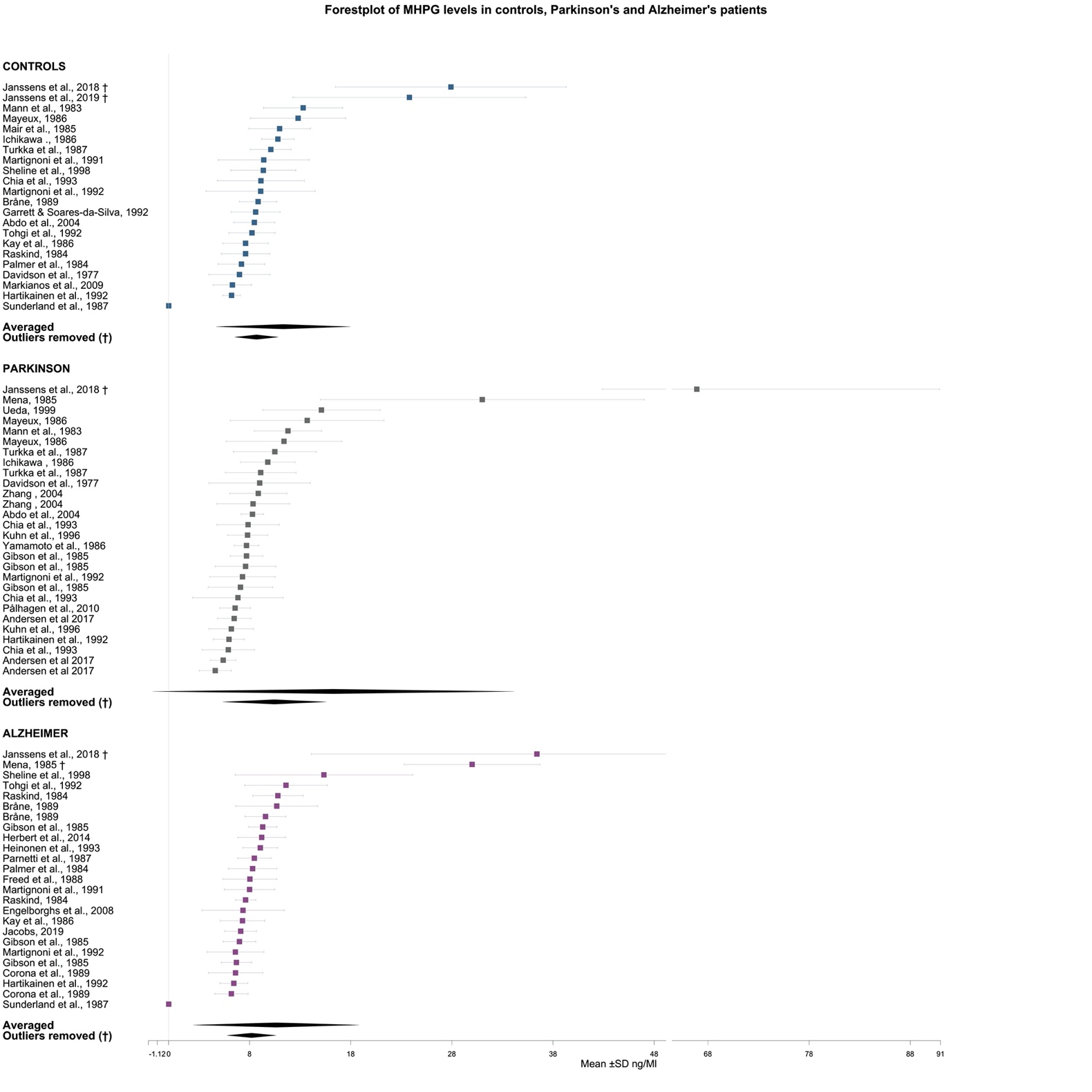

**Supplementary Figure 4.** Forest plot of NA levels (Means and SD) in CSF. Forest plot of means and standard deviations data originated from all articles reporting data points, regardless of their inclusion or exclusion in the meta-analysis. For each study, all datapoints reporting CSF NA or MHPG levels are included. Weighted averaged means of the groups were calculated using the single studies sample sizes as weight. The averages of the weighted means are indicated by the diamonds. CSF: cerebrospinal fluid; NA: noradrenaline; pg/mL: picograms per milliliter.

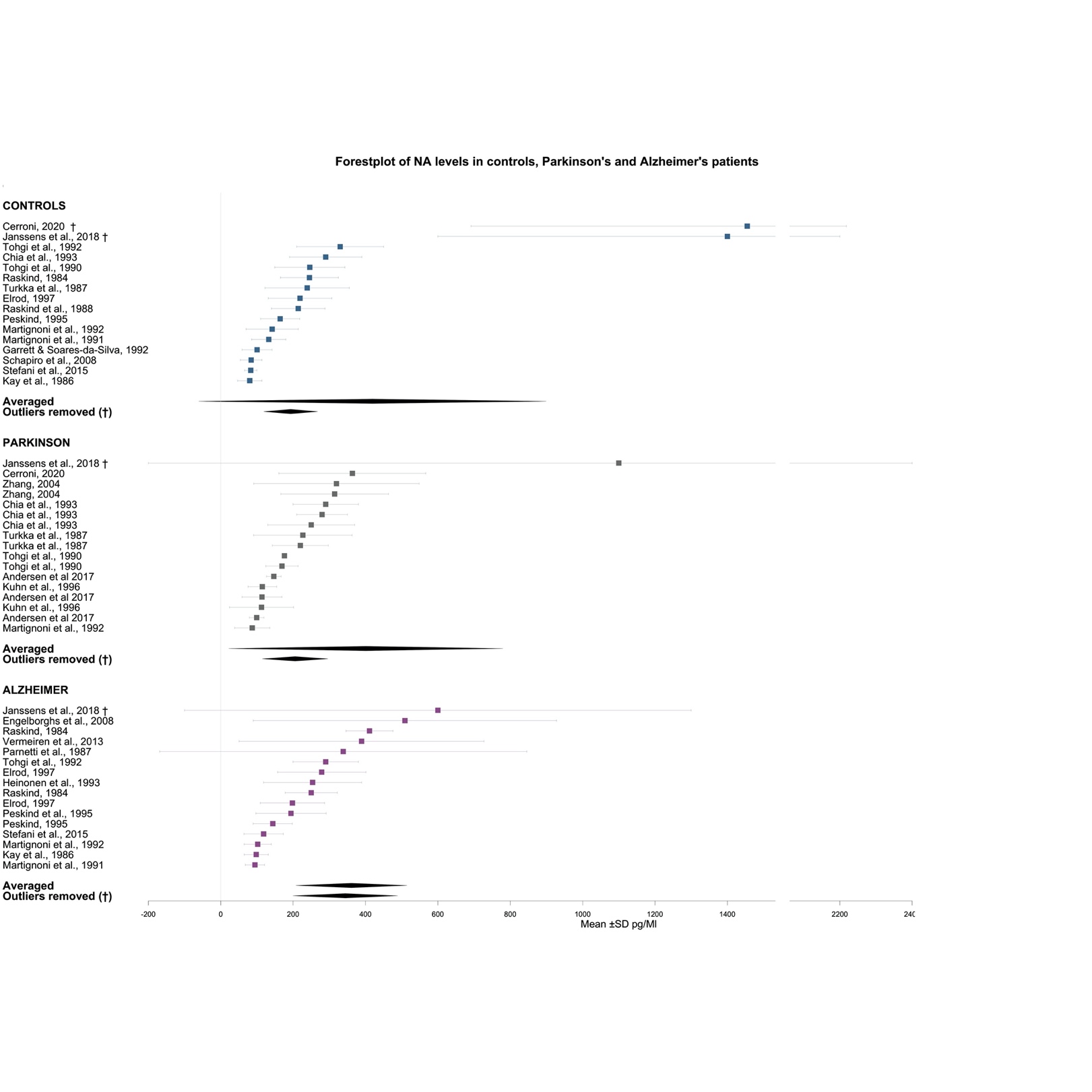

**Supplementary Table 1.** Meta-analysis results. The results are reported before the exclusion of outliers (1), after exclusion based on confidence intervals (2) and after the Leave-One-Out method and the Graphic Display of Heterogeneity plots (3). Between-studies heterogeneity (I-squared) drops consistently in every group after exclusion of studies indicated by the GOSH plot, indicating a decrease in heterogeneity between studies.

| **Group** | **k** | **Hedge’s g** | **se** | **lower** | **upper** | **t** | **p-value** | **I2** | **lower I2** | **upper I2** | **p-value I2** | **out1** | **out2** | **out3** |
| --- | --- | --- | --- | --- | --- | --- | --- | --- | --- | --- | --- | --- | --- | --- |
| NA_ADD 1 | 11 | -0.06 | 0.25 | -0.62 | 0.50 | -0.23 | 0.825 | 78.92% | 0.63 | 0.88 | 0.00 |  |  |  |
| NA_ADD 2 | 11 | -0.06 | 0.25 | -0.62 | 0.50 | -0.23 | 0.825 | 78.92% | 0.63 | 0.88 | 0.00 |  |  |  |
| NA_ADD 3 | 9 | -0.06 | 0.20 | -0.53 | 0.41 | -0.29 | 0.778 | 67.87% | 0.35 | 0.84 | 0.00 | (21) | (22) |  |
| NA_PD 1 | 10 | -0.51 | 0.18 | -0.91 | -0.11 | -2.87 | 0.019 | 45.05% | 0.00 | 0.74 | 0.06 |  |  |  |
| NA_PD 2 | 9 | -0.38 | 0.11 | -0.63 | -0.13 | -3.47 | 0.008 | 0.00% | 0.00 | 0.55 | 0.62 | (23) |  |  |
| NA_PD 3 | 8 | -0.26 | 0.08 | -0.44 | -0.08 | -3.36 | 0.012 | 0.00% | 0.00 | 0.00 | 0.96 | (23) | (13) |  |
| MHPG_ADD 1 | 13 | 0.25 | 0.14 | -0.05 | 0.55 | 1.84 | 0.090 | 45.73% | 0.00 | 0.72 | 0.04 |  |  |  |
| MHPG_ADD 2 | 13 | 0.25 | 0.14 | -0.05 | 0.55 | 1.84 | 0.090 | 45.73% | 0.00 | 0.72 | 0.04 |  |  |  |
| MHPG_ADD 3 | 10 | 0.29 | 0.12 | 0.01 | 0.57 | 2.36 | 0.042 | 11.99% | 0.00 | 0.53 | 0.33 | (15) | (13) | (22) |
| MHPG_PD 1 | 14 | -0.07 | 0.19 | -0.49 | 0.35 | -0.35 | 0.733 | 82.81% | 0.72 | 0.89 | 0.00 |  |  |  |
| MHPG_PD 2 | 13 | -0.27 | 0.08 | -0.44 | -0.09 | -3.37 | 0.006 | 0.00% | 0.00 | 0.18 | 0.90 | (21) |  |  |
| MHPG_PD 3 | 13 | -0.27 | 0.08 | -0.44 | -0.09 | -3.37 | 0.006 | 0.00% | 0.00 | 0.18 | 0.90 | (21) |  |  |

ADD: Alzheimer’s disease dementia; I2= I-squared; High= higher bound of the 95% confidence interval; K= number of studies; Low= lower bound of the 95% confidence interval; MHPG: 3-Methoxy-4-hydroxyphenylglycol; NA: Noradrenaline; PD Parkinson’s disease dementia; se: standard error.

**Supplementary Table 2.** Weighted averages. Weighted averaged means of the groups were calculated using the single studies sample sizes as

weight. The dagger (†) indicates the measures after the exclusion of the outliers.

| **Data** | **Weighted mean** | **Weighted SD** | **N° of articles** | **N° of participants** |
| --- | --- | --- | --- | --- |
| CONTR_MHPG | 11.10 | 6.59 | 22 | 402 |
| PD_MHPG | 16.28 | 17.85 | 28 | 514 |
| ADD_MHPG | 10.63 | 8.18 | 25 | 590 |
| CONTR_NA | 419.13 | 479.95 | 16 | 319 |
| PD_NA | 400.80 | 378.96 | 17 | 229 |
| ADD_NA | 361.03 | 153.53 | 16 | 626 |
| CONTR_MHPG † | 8.66 | 2.14 | 20 | 349 |
| PD_MHPG † | 10.46 | 5.11 | 27 | 461 |
| ADD_MHPG † | 8.19 | 2.43 | 23 | 538 |
| CONTR_NA † | 193.17 | 74.59 | 14 | 260 |
| PD_NA † | 205.49 | 90.88 | 16 | 179 |
| ADD_NA † | 344.28 | 144.70 | 15 | 585 |

ADD: Alzheimer’s disease dementia; CONTR: Controls; MHPG: 3-Methoxy-4-hydroxyphenylglycol; NA: Noradrenaline; PD: Parkinson’s disease dementia; SD: Standard deviation.
